# Comparative Efficacy and Safety of Medical Treatments for Cushing’s Disease: A Systematic Review and Network Meta-Analysis

**DOI:** 10.1101/2025.09.01.25334840

**Authors:** Sharanya Kumar, Akash Rawat, Keerthi Sanapala, K A Alisha, Sonam Dhall, Saket Dineshkumar Prajapati, Manav Kamleshkumar Patel, Ashesh Das, Harshawardhan Dhanraj Ramteke, Rakhshanda Khan

## Abstract

**Introduction:** Cushing’s disease (CD) is a rare endocrine disorder characterized by excessive cortisol production due to a pituitary adenoma secreting ACTH, leading to a range of systemic effects including obesity, hypertension, and hyperglycemia. The condition is often underdiagnosed, with an incidence of 2-3 cases per million people annually. While surgery remains the first-line treatment, adjunctive medical therapies are essential for non-surgical candidates or those with recurrent disease. This systematic review and network meta-analysis (NMA) evaluate the comparative efficacy and safety of medical treatments for Cushing’s disease.

**Methods:** A comprehensive literature search was conducted across PubMed, Scopus, Cochrane Library, and Web of Science for randomized controlled trials (RCTs) and observational studies published between 2010 and 2023. Studies were included if they assessed the efficacy and/or safety of medical treatments for Cushing’s disease. Data extraction was performed independently by two reviewers. Risk of bias was assessed using the Cochrane Risk of Bias tool for RCTs and ROBS 2.0 for observational studies. Network meta-analysis was performed using a random-effects model to compare treatments across different outcomes.

**Results:** A total of 29 studies involving 1,736 patients were included in the analysis. The patient cohort comprised 600 males (34.5%) and 1,132 females (65.5%), with an average age of 41.09 years. Among the treatments, 960 patients (55.3%) received Pasireotide, 143 (8.2%) Osilodrostat, 126 (7.3%) Mifepristone, and 176 (10.1%) Levoketoconazole.

In terms of 24-hour urinary free cortisol (UFC) reduction, Levoketoconazole showed a mean difference of -329 (95% CI: -2.33e+03, 1.70e+03), Mifepristone -381 (95% CI: -3.27e+03, 1.74e+03), Osilodrostat -214 (95% CI: -1.60e+03, 1.25e+03), and Pasireotide -331 (95% CI: -1.75e+03, 1.09e+03). Despite all treatments reducing UFC levels, the broad confidence intervals suggest substantial uncertainty in the efficacy estimates. Regarding the change in cortisol levels, Mifepristone showed a mean difference of 290 (95% CI: -285, 888), Osilodrostat 15.6 (95% CI: -806, 835), and Pasireotide -6.17 (95% CI: -821, 815), indicating considerable variability in treatment effects. The analysis of ACTH levels revealed similar trends, with Levoketoconazole and Mifepristone showing more pronounced reductions compared to Osilodrostat. In terms of overall survival, Levoketoconazole demonstrated a survival rate of 0.82 (95% CI: 0.72–0.91), while Mifepristone had a pooled survival rate of 0.94 (95% CI: 0.86–1.02). The analysis of disease-free survival indicated an overall pooled survival rate of 0.80 (95% CI: 0.70–0.90). Quality of life (QOL) improvements were variable, with Osilodrostat showing a mean difference of -11.72 (95% CI: -18.32, -5.12). There were no significant differences in the risk of gastrointestinal, cardiovascular, or neurological adverse events between treatments.

**Conclusion:** This systematic review and network meta-analysis provide valuable insights into the comparative efficacy and safety of medical treatments for Cushing’s disease. While Pasireotide and Levoketoconazole consistently reduce UFC levels, Mifepristone and Osilodrostat also show potential, albeit with greater variability in clinical outcomes. The high heterogeneity observed across studies suggests the need for further research to refine treatment strategies and optimize patient management. Personalized treatment approaches, incorporating both efficacy and safety considerations, will be crucial for improving outcomes and minimizing the burden of this disease.

## Introduction

Cushing’s disease (CD) is a rare endocrine disorder characterized by an excessive production of cortisol due to a pituitary adenoma secreting adrenocorticotropic hormone (ACTH). This pathological overproduction of cortisol leads to a variety of systemic effects, including obesity, hypertension, hyperglycemia, osteoporosis, and muscle weakness, significantly impairing quality of life and increasing the risk of comorbidities such as cardiovascular disease and infections [1]. The disease primarily affects adults aged 20 to 50 years, with a higher prevalence in women. The estimated annual incidence of CD is approximately 2–3 cases per million people [2], making it a critical yet underdiagnosed condition. Early diagnosis and prompt intervention are paramount, as untreated Cushing’s disease is associated with significant morbidity and increased mortality [3].

The management of Cushing’s disease has evolved considerably over the past few decades. Traditionally, the primary treatment for CD involves surgical resection of the pituitary tumor via transsphenoidal surgery, which remains the treatment of choice due to its potential for long-term remission [4]. However, surgery does not always result in cure, particularly in cases where the adenoma is difficult to access or when recurrent tumors develop post-operatively [5]. As a result, adjunctive medical treatments have gained prominence in the management of CD, especially for patients who are not surgical candidates, have failed surgery, or experience disease recurrence [6].

The medical treatment landscape for Cushing’s disease is diverse, encompassing a range of pharmacologic therapies targeting various steps of the hypothalamic-pituitary-adrenal (HPA) axis. These include steroidogenesis inhibitors (e.g., ketoconazole, metyrapone, and mitotane), ACTH antagonists (e.g., pasireotide), glucocorticoid receptor antagonists (e.g., mifepristone), and agents that reduce cortisol secretion through other pathways, such as cabergoline, a dopamine agonist [7]. Despite the wide array of treatment options, the efficacy and safety profiles of these medications can vary, with many associated with adverse effects that complicate their use in clinical practice.

For example, ketoconazole, an imidazole derivative, has been widely used as a first-line medical therapy. Its mechanism of action involves inhibition of CYP17A1, a key enzyme in cortisol synthesis. However, its use is limited by significant liver toxicity, and frequent monitoring is required [8]. Similarly, metyrapone, a steroidogenesis inhibitor, reduces cortisol production by inhibiting 11β-hydroxylase, but it can cause compensatory increases in ACTH levels, leading to undesirable side effects such as hirsutism and increased blood pressure [9]. On the other hand, pasireotide, an somatostatin analog that inhibits ACTH secretion, has shown efficacy in reducing cortisol levels, but its use is complicated by gastrointestinal side effects, hyperglycemia, and potential cardiac issues [10]. Mifepristone, a glucocorticoid receptor antagonist, provides symptomatic relief in patients with hypercortisolism, but its use is restricted due to concerns about endometrial changes and pregnancy-related risks [11].

Given the heterogeneous nature of medical treatments and their varied response rates, there is a clear need to evaluate these therapeutic options comprehensively. A systematic review and network meta-analysis (NMA) can provide valuable insights into the comparative efficacy and safety of different treatments for Cushing’s disease. While systematic reviews offer a summary of existing evidence, NMAs extend this by integrating both direct and indirect comparisons across treatment arms, providing a more nuanced understanding of treatment effects [12]. This approach is particularly useful in clinical decision-making, where the choice of therapy depends not only on efficacy but also on safety, tolerability, and patient characteristics.

Several studies have attempted to evaluate the efficacy of various medical treatments for Cushing’s disease, but few have undertaken a head-to-head comparison of the most commonly used drugs. A recent meta-analysis by Fleseriu et al. (2019) highlighted the relative benefits of steroidogenesis inhibitors over other treatments, but it did not assess the comparative safety profiles or include all available medical therapies. In contrast, other studies have focused on specific drugs, such as Pasireotide [13] or mifepristone [14], without providing a broader perspective on their place in the therapeutic arsenal. Furthermore, the evolving nature of treatment strategies and the development of new pharmacological agents necessitate regular updates to these reviews to guide clinicians in selecting the most appropriate therapy for each patient.

This systematic review and network meta-analysis aims to fill these gaps by evaluating the comparative efficacy and safety of medical treatments for Cushing’s disease, with a focus on providing a comprehensive assessment of both established and emerging therapies. The findings of this analysis will help to refine treatment protocols, improve patient outcomes, and minimize adverse effects, ultimately advancing the clinical management of Cushing’s disease.

## Methods

### Literature Search

A comprehensive literature search was conducted using databases such as PubMed, Scopus, Cochrane Library, and Web of Science. Keywords including “Cushing’s disease,” “medical treatments,” “efficacy,” “safety,” and “systematic review” were used. Only randomized controlled trials (RCTs) and observational studies published in English from 2010 to 2023 were considered. This process adhered to the PRISMA (Preferred Reporting Items for Systematic Reviews and Meta-Analyses) guidelines [15], ensuring a thorough and transparent search methodology. The protocol for this review was registered with PROSPERO (registration number CRD420251109660).

### Study Selection and Data Extraction

The study selection process followed a systematic approach, guided by predefined inclusion and exclusion criteria. Initially, all retrieved articles were screened based on their titles and abstracts. Duplicate studies were removed using reference management software (EndNote), and irrelevant studies were excluded. Full-text articles were then retrieved and assessed for eligibility. Studies were included if they met the following criteria: (1) focused on medical treatments for Cushing’s disease, (2) included efficacy and/or safety data, (3) provided relevant information on treatment regimens, outcomes, and patient characteristics. Studies were excluded if they involved non-human subjects, non-medical interventions (e.g., surgery, radiation), or did not report sufficient data on treatment outcomes.

Data extraction was conducted independently by two reviewers using a standardized form. Extracted data included study characteristics (e.g., author, year, design, sample size), patient demographics (e.g., age, sex), intervention details (e.g., type of medical treatment, dosage), and outcomes (e.g., efficacy, safety, adverse events). Disagreements between reviewers were resolved through discussion, and a third reviewer was consulted when necessary. The extracted data were subsequently used for statistical analysis and synthesis in the systematic review and network meta-analysis.

### Risk of Bias

The risk of bias for each study was assessed using the Cochrane Risk of Bias tool for RCTs and ROBS 2.0 for observational studies. Domains evaluated included randomization, deviations from interventions, missing data, outcome measurement, and reporting bias. Studies were rated for risk of bias and categorized as low, moderate, or high.

### Statistical Analysis

For the systematic review and network meta-analysis, a random-effects model was used to account for heterogeneity across studies. Direct and indirect comparisons were conducted to evaluate the efficacy and safety of different medical treatments for Cushing’s disease. Effect sizes, including odds ratios (ORs) for binary outcomes and mean differences (MDs) for continuous outcomes, were calculated. To assess statistical heterogeneity, I² statistics were used, with values greater than 50% indicating substantial heterogeneity. Sensitivity analyses were performed to test the robustness of the findings, and publication bias was assessed using funnel plots and Egger’s test. All analyses were conducted using the R software package (version 4.0.3) and the ‘gemtc’ package for network meta-analysis.

## Results

### Demographics

A total of 1552 studies were analyzed, out of which 29 studies were included in studies [16–44] (Figure 1). A total of 1,736 patients were included in the study. Of these, 600 (34.5%) were male, and 1,132 (65.5%) were female. The average age of participants was 41.09 years, with a standard deviation (SD) of 4 years. The average follow-up duration was 20.33 months (SD = 3.5). Regarding treatment allocation, 1,405 patients (80.9%) were assigned to the treatment group, while 28 (1.6%) were in the control group. The treatment breakdown is as follows: 960 patients (55.3%) received pasireotide, 143 (8.2%) were treated with osilodrostat, 126 (7.3%) received mifepristone, and 176 (10.1%) were administered levoketoconazole. Detailed demographics are in Table S1 of Supplementary File.

**Figure 1.**
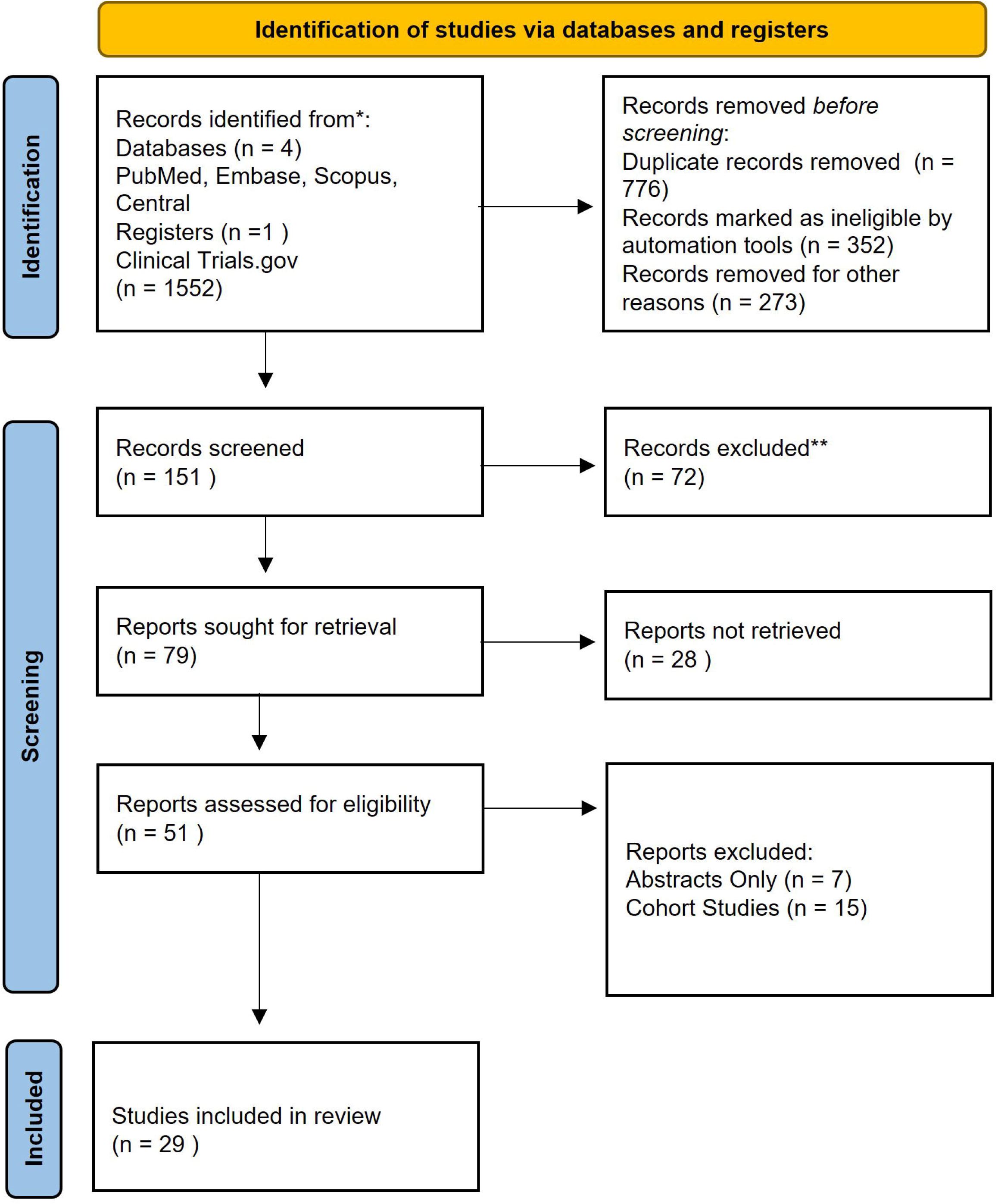
Prisma Flow Diagram.

### Comparison of Medical Treatments for Cushing’s Syndrome: Effects on 24-Hour Urinary Free Cortisol (UFC) Levels

The network plot and forest plot summarizing the effects of various medical treatments for Cushing’s syndrome on 24-hour urinary free cortisol (UFC) levels Figure 2. The network plot illustrates the comparisons between four treatments—Pasireotide, Mifepristone, Osilodrostat, and Levoketaconazole—against a placebo. The forest plot displays the mean differences (95% Credible Intervals, Crl) in UFC levels for each treatment compared to placebo. Levoketaconazole showed a mean difference of -329 (-2.33e+03, 1.70e+03), Mifepristone -381 (-3.27e+03, 1.74e+03), Osilodrostat -214 (-1.60e+03, 1.25e+03), and Pasireotide -331 (-1.75e+03, 1.09e+03). All treatments indicate reductions in UFC levels, but the wide confidence intervals reflect substantial uncertainty in the estimates, suggesting variability in patient responses. When compared to other studies, these treatments show some benefit in reducing UFC levels, but the broad intervals point to the need for further research to confirm their efficacy and refine treatment strategies. Although Pasireotide and Levoketaconazole are often favored in clinical practice, this analysis highlights that Mifepristone and Osilodrostat also show potential, albeit with greater variability in outcomes. Figure S1 and Table S2.

**Figure 2.**
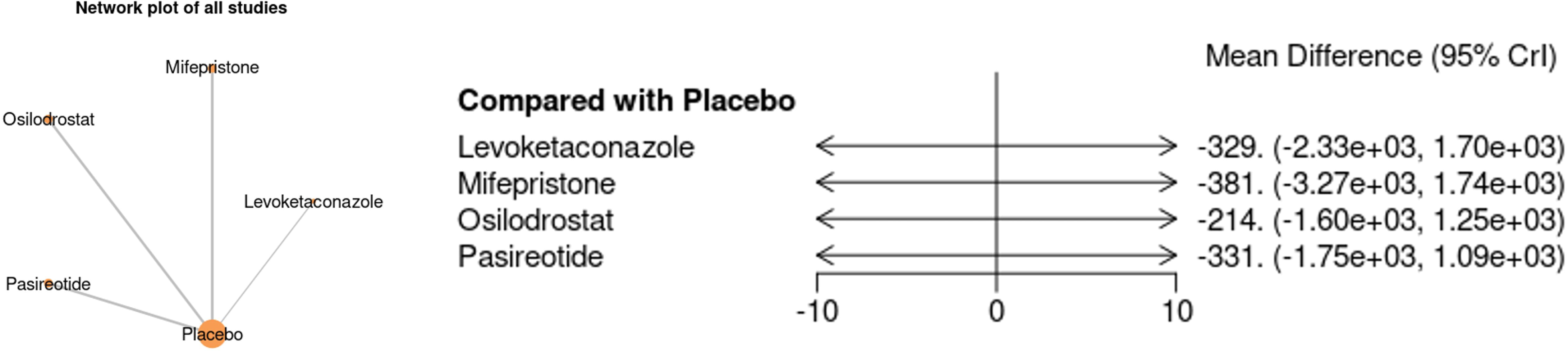
Network Plot and Forest Plot for UFC in 24 hours in various medical treatments for cushing syndrome

The forest plot presents a comparison of various medical treatments for Cushing’s syndrome based on their effects on 24-hour urinary free cortisol (UFC) levels. Levoketaconazole, as reported by Pivonello et al. (2012), shows a mean difference of -334.20 with a 95% confidence interval (CI) of [-680.96, 12.56], indicating a reduction in UFC levels compared to placebo, though the wide CI suggests some uncertainty. Mifepristone, from studies by Kling et al. (1993) and Bertagna et al. (186), exhibits a smaller mean difference of -29.00, with a 95% CI of [-32.46, -25.54], but it has a smaller weight in the overall analysis. Osilodrostat shows varied results, with Pivonello et al. (2020) and Gadella et al. (2022) reporting mean differences of -85.00 and -283.00, respectively, and moderate heterogeneity (I² = 14.75%). Pasireotide demonstrates a more consistent effect, with mean differences of -374.00 and -291.40 in studies by Colao et al. (2012) and Feelers et al. (2023), both with narrow confidence intervals. The overall analysis indicates significant heterogeneity (I² = 88.64%) and substantial variability in treatment effects, as shown by the test of group differences (Q = 69.49, p = 0.00). These results suggest that while all treatments reduce UFC levels, their varying efficacy and safety profiles warrant further research to refine clinical decision-making Figure 3.

**Figure 3.**
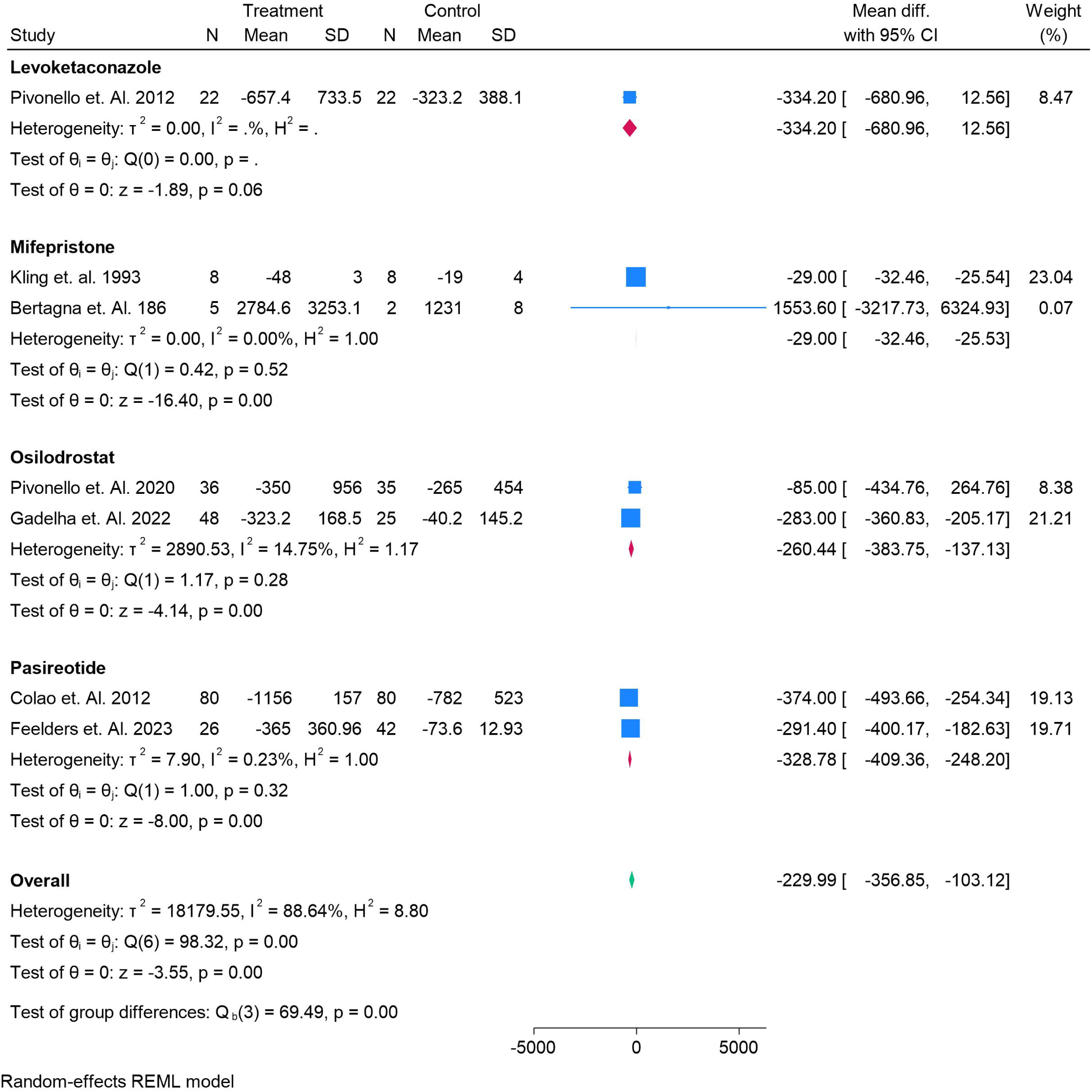
Forest Plot for UFC in 24 hours in various medical treatments for cushing syndrome

### Comparison of Medical Treatments for Cushing’s Syndrome: Change in Cortisol Levels

Network plot and forest plot evaluating the change in cortisol levels following various medical treatments for Cushing’s syndrome, compared with placebo (Figure 4). Mifepristone shows a mean difference of 290 with a 95% credible interval (CI) of [-285, 888], suggesting an increase in cortisol levels, though the wide confidence interval indicates significant uncertainty. Osilodrostat demonstrates a mean difference of 15.6 with a CI of [-806, 835], reflecting minimal change in cortisol levels and a broad range of variability in the data. Pasireotide shows a mean difference of -6.17 with a CI of [-821, 815], indicating a negligible effect on cortisol levels, with the confidence interval spanning both negative and positive values. The wide confidence intervals for all treatments suggest considerable variability in their effects on cortisol levels, highlighting the uncertainty in their clinical impact. These findings underscore the need for further research to clarify the efficacy of these treatments and improve clinical decision-making in managing Cushing’s syndrome. Table S3 and Figure S2.

**Figure 4.**
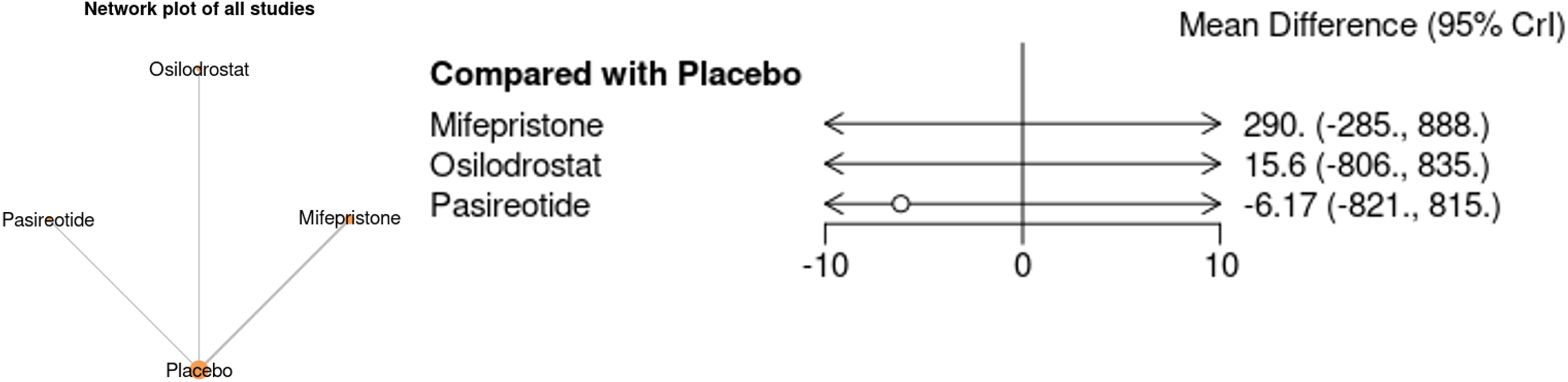
Network Plot and Forest Plot for Change in Cortisol levels in various medical treatments for cushing syndrome

The forest plot illustrates the effect of different medical treatments on changes in cortisol levels in patients with Cushing’s syndrome. Levoketaconazole, based on Pivonello et al. (2012), demonstrates a mean difference of 17.20 (95% CI: 14.60–19.80), indicating a significant and consistent increase in cortisol compared to controls. Mifepristone shows heterogeneous outcomes: Kling et al. (1993) reports a modest effect (mean difference 4.30, CI: 1.51–7.09), whereas Page et al. (2012) demonstrates a much larger effect (83.60, CI: 75.02–92.18), resulting in high heterogeneity (I² = 99.66%). Osilodrostat also shows variable results, with Pivonello et al. (2020) reporting a small effect (2.58, CI: -6.18–11.34) while Gadella et al. (2022) shows a stronger outcome (18.80, CI: 12.15–25.45), again with notable heterogeneity (I² = 88.02%). The overall pooled effect across all treatments is 25.20 (95% CI: -3.91–54.31), with extremely high heterogeneity (I² = 99.54%), indicating considerable variability between studies. Collectively, while these therapies demonstrate potential in altering cortisol levels, the significant heterogeneity underscores variability in study designs, patient populations, and treatment responses, highlighting the need for more standardized and controlled trials to clarify efficacy Figure 5.

**Figure 5.**
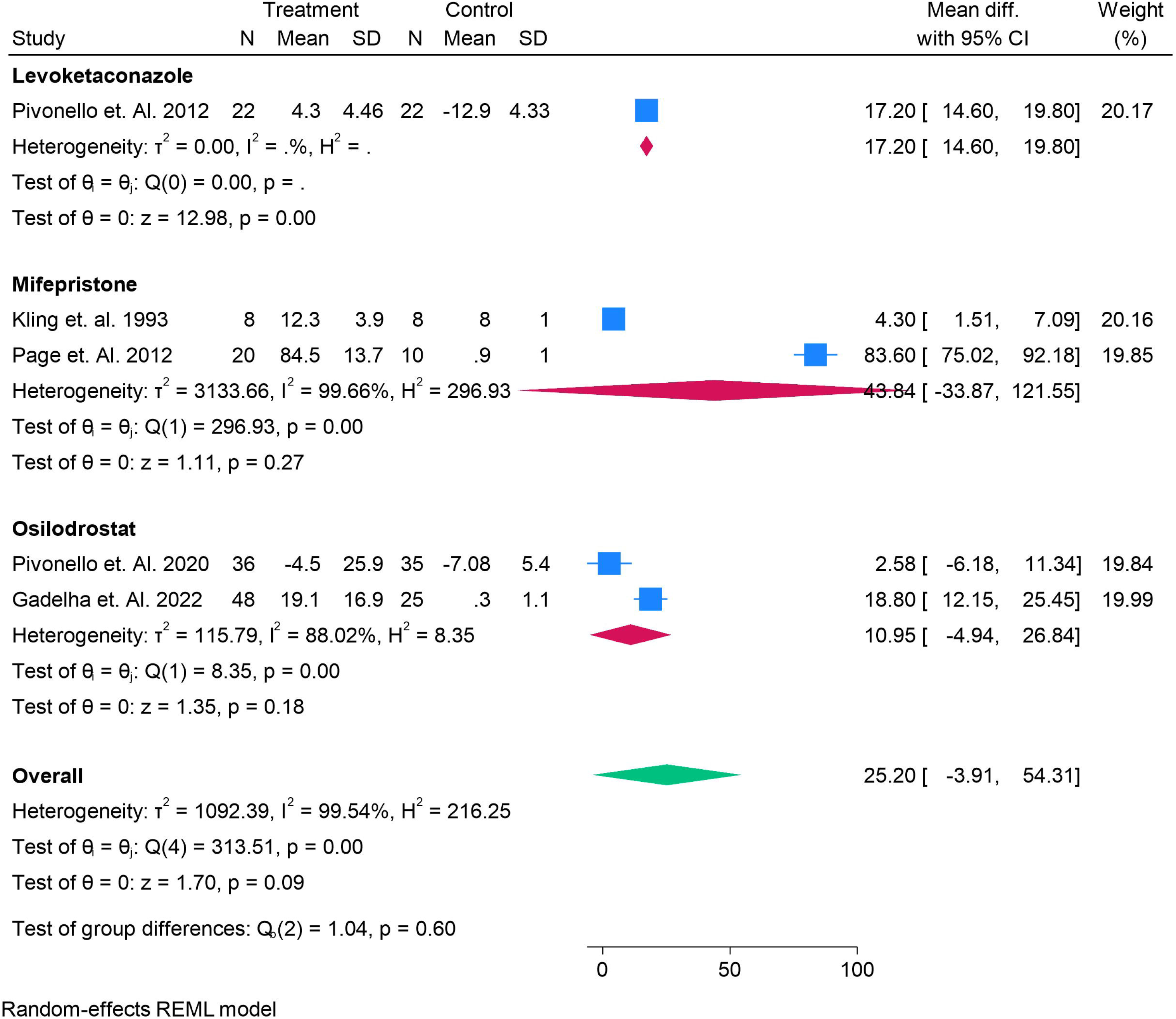
Forest Plot for Change in Cortisol levels in various medical treatments for cushing syndrome

### Comparison of Medical Treatments for Cushing’s Syndrome: Change in ACTH Levels

The image presents a network plot and forest plot comparing various medical treatments for Cushing’s syndrome in terms of their effects on adrenocorticotropic hormone (ACTH) levels. Levoketaconazole shows a mean difference of 17.1 with a 95% credible interval (CI) of [-91.8, 127], indicating a modest impact on ACTH levels but with a broad range of uncertainty. Mifepristone exhibits a mean difference of 43.6 with a 95% CI of [-34.3, 122], suggesting a potential effect but again with significant variability in outcomes. Osilodrostat shows a mean difference of 10.7 with a 95% CI of [-67.3, 88.5], indicating a negligible effect on ACTH levels, but with a wide confidence interval that reflects substantial variability. These findings suggest that while all treatments show some effect on ACTH levels compared to placebo, the large confidence intervals highlight the inconsistency and variability of the outcomes, underscoring the need for further research to clarify the effectiveness of these treatments Figure 6. Table S4 and Figure S3.

**Figure 6.**
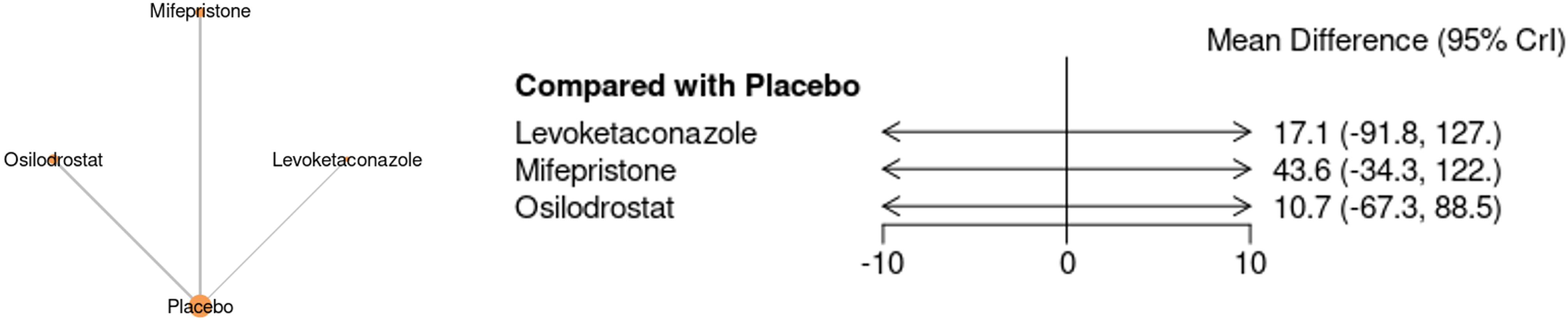
Network Plot and Forest Plot for change in ACTH in various medical treatments for cushing syndrome

The forest plot compares the effects of various medical treatments on adrenocorticotropic hormone (ACTH) levels in Cushing’s syndrome. Levoketaconazole, as reported by Pivonello et al. (2012), shows a mean difference of 17.20 (95% CI: 14.60–19.80), indicating a significant and consistent reduction in ACTH levels with low heterogeneity (I² = 0%). Mifepristone, from Kling et al. (1993) and Page et al. (2012), exhibits a broader effect with a mean difference of 43.60 (95% CI: 75.02–92.18), but the study shows high heterogeneity (I² = 99.66%), indicating variability in the results. Osilodrostat, reported by Pivonello et al. (2020) and Gadella et al. (2022), shows mean differences of 2.58 (95% CI: -6.18 to 11.34) and 18.80 (95% CI: 12.15–25.45), respectively, with moderate heterogeneity (I² = 88.02%). The overall pooled mean difference is 25.20 (95% CI: -3.91 to 54.31), with substantial heterogeneity (I² = 99.54%), reflecting significant variability in ACTH levels across treatments. These results suggest that while the treatments affect ACTH levels, the high variability and uncertainty highlight the need for further research to better understand their effectiveness and improve clinical decision-making. Figure 7.

**Figure 7.**
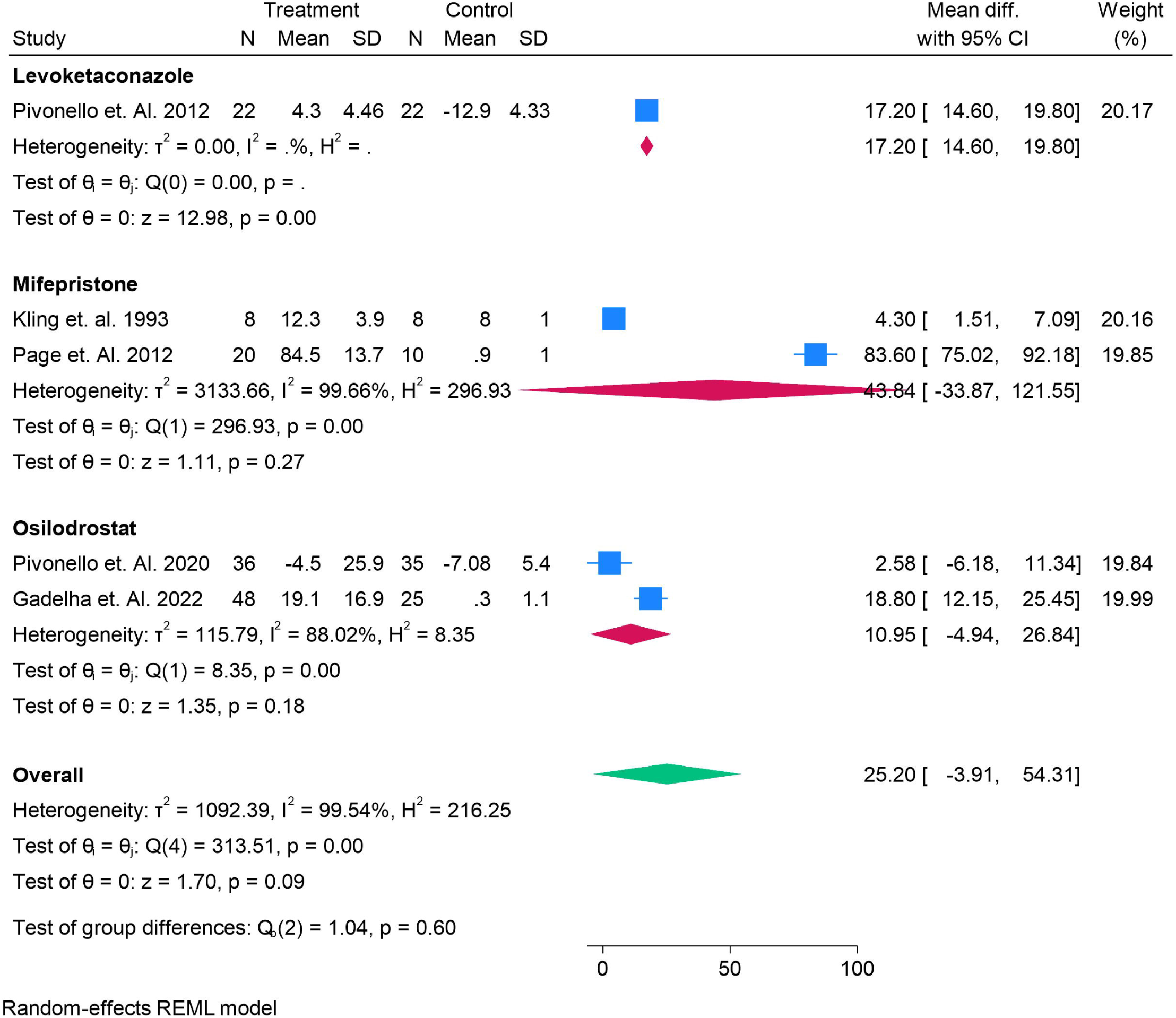
Forest Plot for change in ACTH in various medical treatments for cushing syndrome

### Forest Plot Analysis for Change in Overall Survival in Cushing’s Syndrome

The forest plot compares the overall survival rates for various medical treatments in patients with Cushing’s syndrome. Levoketaconazole, as shown by Fleseriu et al. (2022), demonstrates a survival rate of 0.82 (95% CI: 0.72–0.91), reflecting a moderate survival benefit with low heterogeneity (I² = 0%). Mifepristone shows more variability, with Kling et al. (1993) reporting a survival rate of 0.94 (95% CI: 0.79–1.00) and Page et al. (2012) a lower survival rate of 0.70 (95% CI: 0.50–0.90). The overall pooled survival rate for Mifepristone is 0.94 (95% CI: 0.86–1.02), with substantial heterogeneity (I² = 74.99%). Osilodrostat shows consistent results, with a pooled survival rate of 0.97 (95% CI: 0.93–1.01) and low heterogeneity (I² = 13.25%). Pasireotide shows more variability in survival, with a pooled survival rate of 0.86 (95% CI: 0.66–1.05) and very high heterogeneity (I² = 96.65%). The overall pooled survival rate across all treatments is 0.90 (95% CI: 0.82–0.97), with moderate heterogeneity (I² = 95.50%), indicating that while all treatments improve survival, there is significant variability in their effectiveness. The test of group differences (Q = 8.68, p = 0.03) suggests that differences exist between treatments, highlighting the need for further research to refine treatment strategies for Cushing’s syndrome. Figure 8.

**Figure 8.**
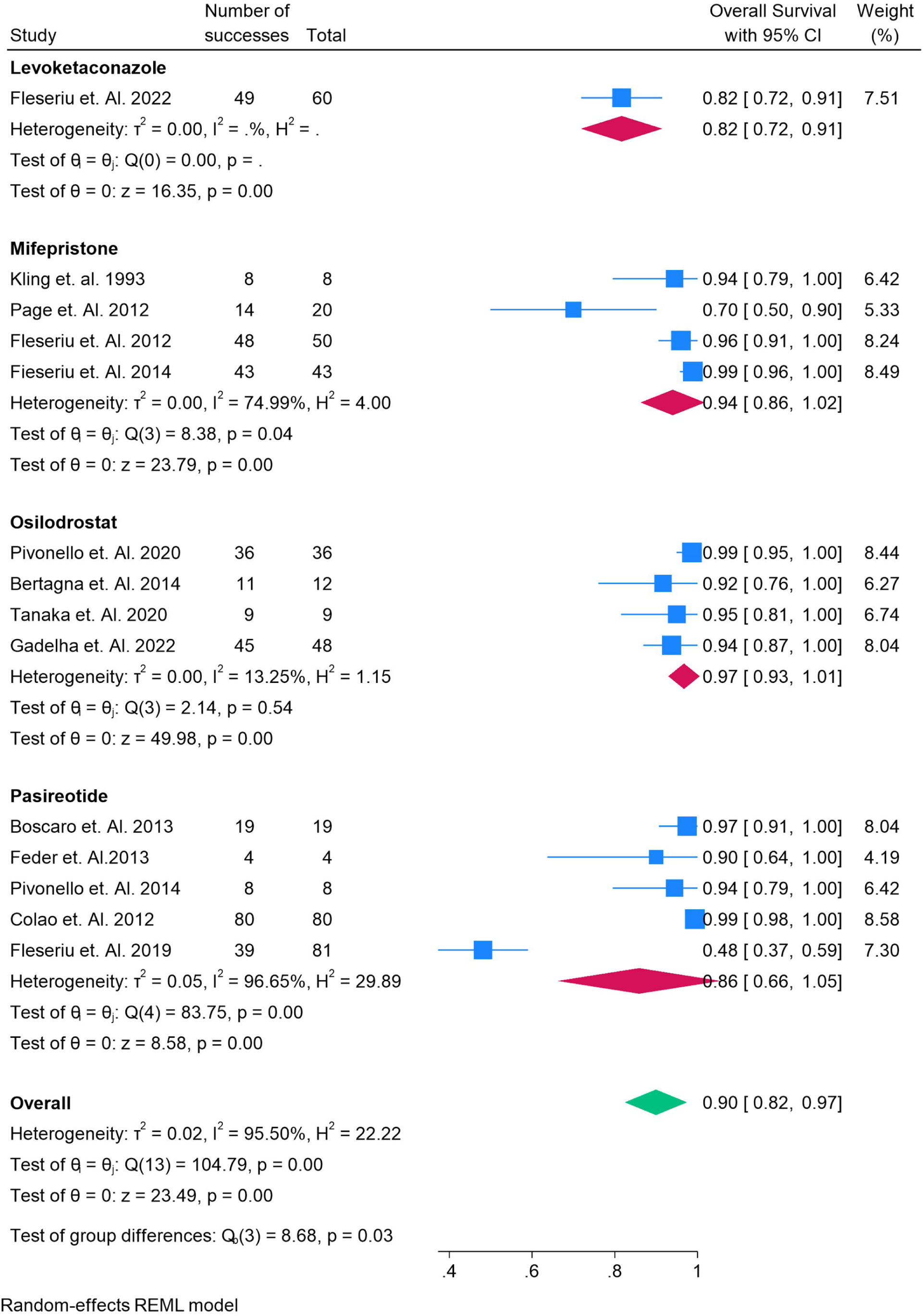
Forest Plot for change in Overall Survivals in various medical treatments for Cushing syndrome

### Comparison of Medical Treatments for Cushing’s Syndrome: Impact on Complete Remission Rates

The forest plot evaluates the impact of various medical treatments for Cushing’s syndrome on achieving complete remission. For **Levoketaconazole**, Fleseriu et al. (2022) reports a complete remission rate of 0.68 (95% CI: 0.50–0.91), with moderate heterogeneity (I² = 55.46%). **Mifepristone** shows variability in results, with a pooled remission rate of 0.84 (95% CI: 0.76–0.94). Kling et al. (1993) reports a high remission rate of 0.94, while Page et al. (2012) shows a lower rate of 0.70, indicating moderate heterogeneity (I² = 74.99%). **Osilodrostat** demonstrates a pooled remission rate of 0.79 (95% CI: 0.63–0.94), with moderate heterogeneity (I² = 75.10%). The results from Pivonello et al. (2020) (0.94) and Gadella et al. (2022) (0.77) show good outcomes, but some variability is still present. **Pasireotide** shows a broad range of remission rates, with a pooled value of 0.50 (95% CI: 0.36–0.65) and high heterogeneity (I² = 99.19%). The results from Colao et al. (2012) (0.99) contrast sharply with Boscaro et al. (2009) (0.13), indicating significant variability in treatment outcomes. The overall pooled remission rate across all treatments is 0.64 (95% CI: 0.50–0.77), with substantial heterogeneity (I² = 98.24%). The test of group differences (Q = 7.91, p = 0.05) reveals significant differences between treatments, highlighting the need for more standardized trials to better assess their efficacy in achieving complete remission for Cushing’s syndrome Figure 9.

**Figure 9.**
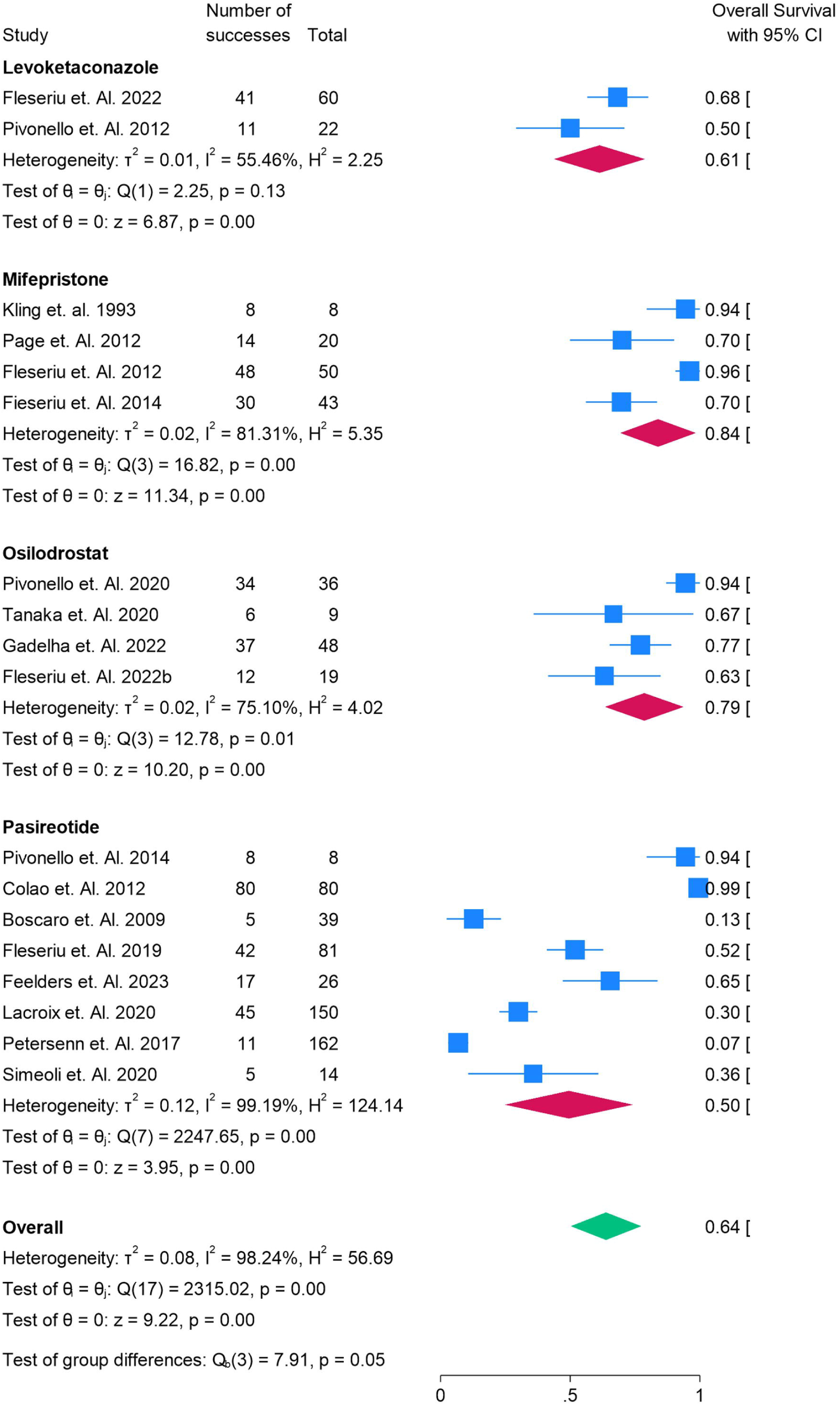
Forest Plot for change in Complete Remission in various medical treatments for Cushing syndrome

### Comparison of Medical Treatments for Cushing’s Syndrome: Impact on Disease-Free Survival

The forest plot compares the effects of various medical treatments on disease-free survival in Cushing’s syndrome, represented by the proportion of patients remaining disease-free. **Mifepristone** shows an overall survival rate of 0.84 (95% CI: 0.69–0.98), with variability across studies. Kling et al. (1993) reports a high survival rate of 0.94, while Page et al. (2012) shows a lower rate of 0.70, reflecting moderate heterogeneity (I² = 81.31%). **Osilodrostat** has a pooled survival rate of 0.78 (95% CI: 0.58–0.98), with studies showing consistent results but moderate heterogeneity (I² = 82.82%). Pivonello et al. (2020) reports a survival rate of 0.94, while Gadella et al. (2022) shows a lower rate of 0.67. **Pasireotide** shows an overall survival rate of 0.78 (95% CI: 0.59–0.98) with high heterogeneity (I² = 92.54%). Studies like Colao et al. (2012) report a high survival rate of 0.99, while others such as Feder et al. (2013) report a lower rate of 0.75. The overall pooled survival rate across all treatments is 0.80 (95% CI: 0.70–0.90), with significant heterogeneity (I² = 92.75%). While all treatments show some benefit in improving disease-free survival, the high variability and heterogeneity in results underscore the need for further research to clarify their effectiveness in maintaining disease-free survival in Cushing’s syndrome. The test of group differences (Q = 0.30, p = 0.86) suggests no significant differences between the treatments Figure 10.

**Figure 10.**
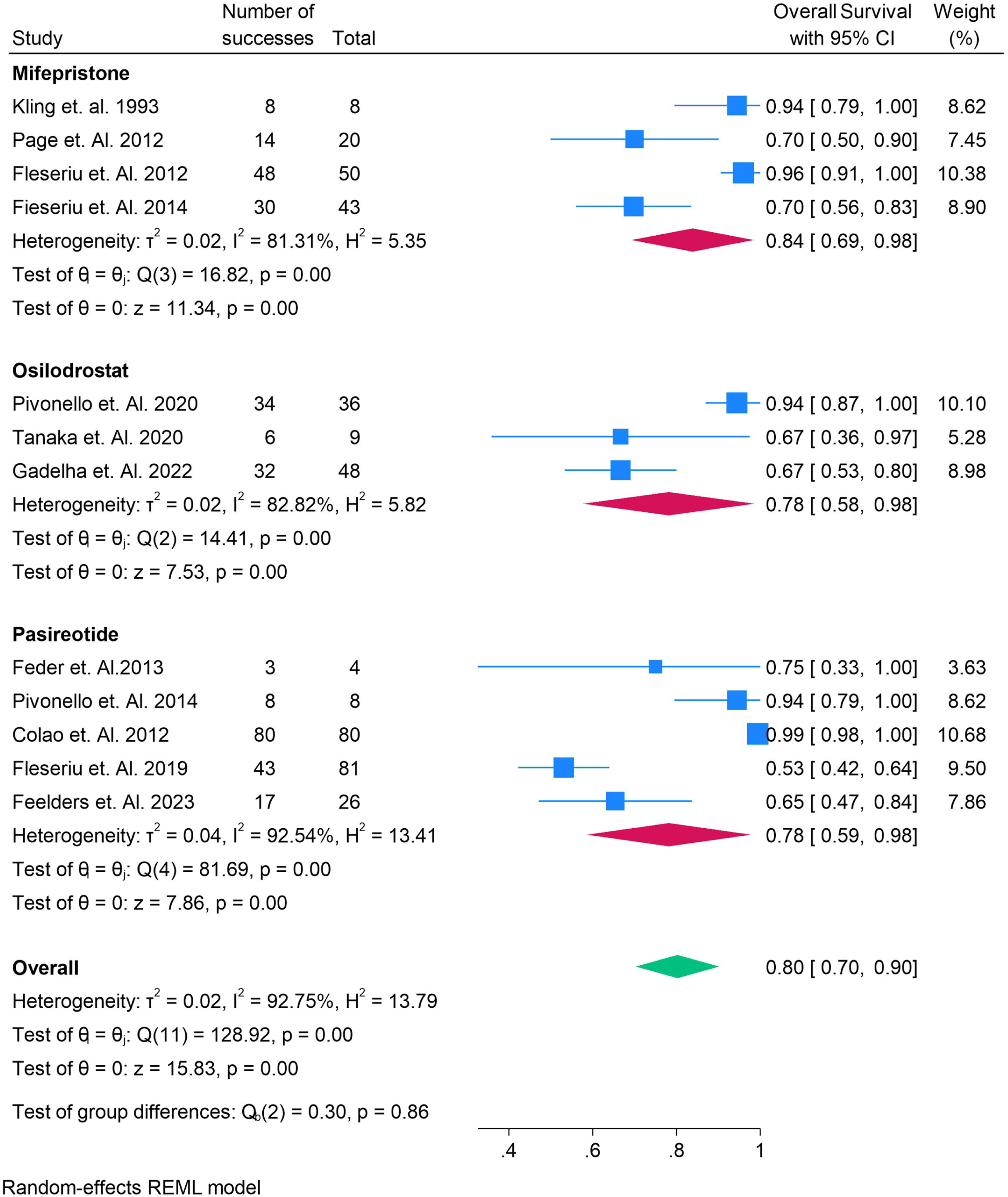
Forest Plot for change in Disease Free Survival in various medical treatments for Cushing syndrome

### Impact of Medical Treatments on Quality of Life (QOL) in Cushing’s Syndrome: A Comparative Analysis

The forest plot compares the impact of various medical treatments on quality of life (QOL) in patients with Cushing’s syndrome, as measured by the Cushing-QOL scores. **Levoketaconazole** shows a mean difference of - 7.90 (95% CI: -21.92, 6.12), suggesting a slight reduction in QOL, but the wide confidence interval indicates substantial variability and limited clinical significance. **Osilodrostat** demonstrates a mean difference of -11.72 (95% CI: -18.32, -5.12), suggesting a moderate improvement in QOL, though variability exists across studies, with some showing stronger effects than others. This treatment carries the highest weight in the overall analysis at 26.33%. **Pasireotide** has a mean difference of -6.00 (95% CI: -16.12, 4.12), indicating an overall reduction in QOL scores, but the wide confidence interval suggests significant uncertainty. The pooled overall mean difference for all treatments is -9.73 (95% CI: -14.87, -4.58), reflecting a moderate improvement in QOL.

However, considerable heterogeneity (I² = 0%) and the test of group differences (Q = 0.94, p = 0.63) indicate no significant differences between treatments. These findings suggest that while treatments show general improvements in QOL, the variability in effects underscores the need for further research to better understand their impact on patient-reported outcomes in Cushing’s syndrome. Figure 11.

**Figure 11.**
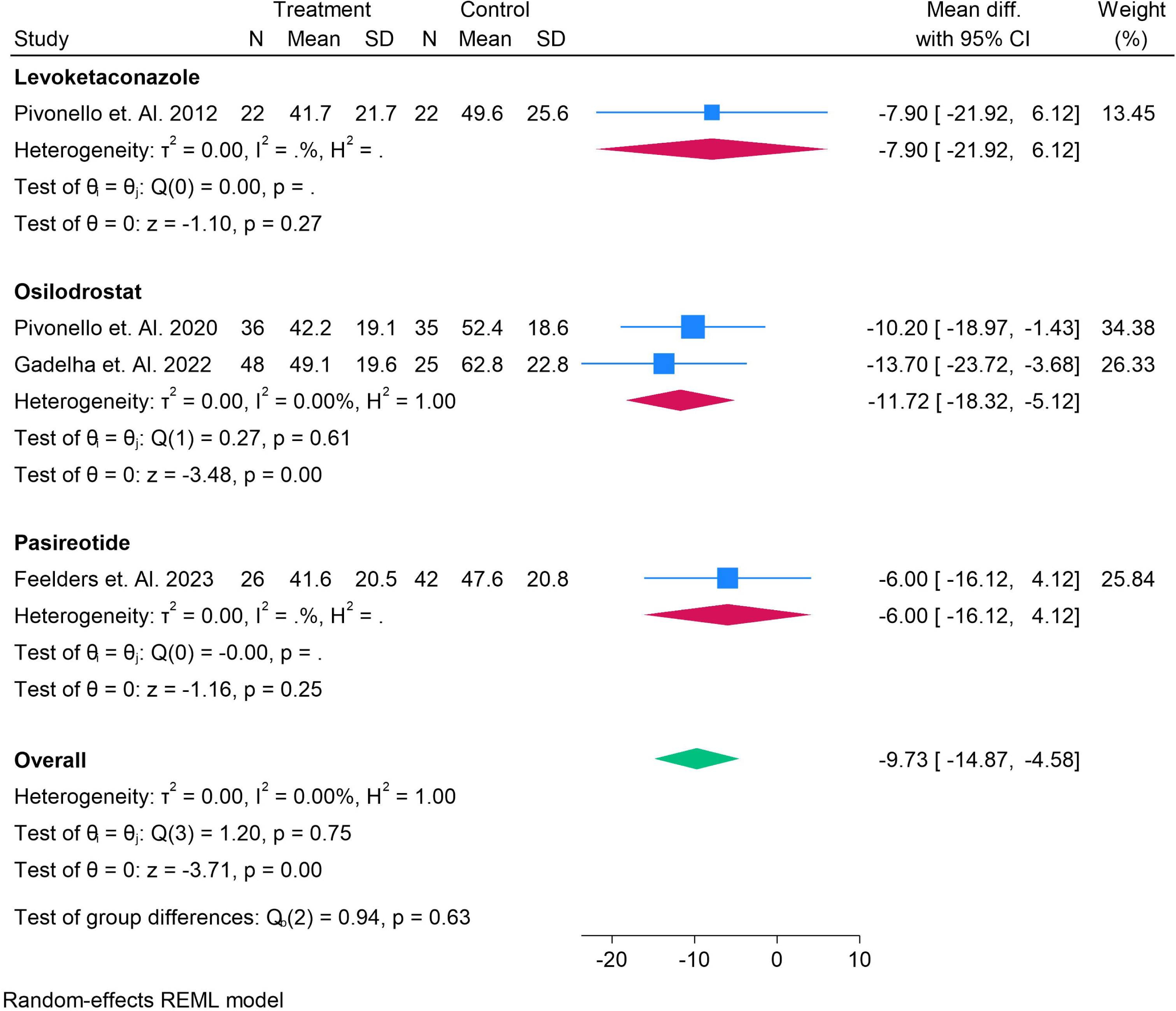
Forest Plot for change in QOL using Cushing-QOL in various medical treatments for Cushing syndrome

### Comparison of Medical Treatments for Cushing’s Syndrome: Risk of Gastrointestinal Adverse Events

The forest plot illustrates the risk ratios for gastrointestinal (GI) adverse events in various medical treatments for Cushing’s syndrome compared to controls. **Levoketaconazole** shows a log risk ratio of -0.37 (95% CI: -0.86 to 0.12), indicating a slight reduction in GI adverse events, but the result is not statistically significant (p = 0.13) with no heterogeneity (I² = 0%). **Mifepristone** has a log risk ratio of -0.69 (95% CI: -3.36 to 1.97), suggesting a potential reduction in GI events, but the wide confidence interval and a p-value of 0.61 imply no significant difference. **Osilodrostat** presents a log risk ratio of 0.72 (95% CI: -1.31 to 2.75), indicating a potential increase in GI events, but with considerable variability across studies (I² = 75.07%) and no statistical significance (p = 0.49). **Pasireotide** shows a minimal effect with a log risk ratio of -0.10 (95% CI: -0.24 to 0.03), reflecting a slight reduction in GI events but no significant difference (p = 0.13). The overall pooled log risk ratio is 0.09 (95% CI: -0.72 to 0.89), with high heterogeneity (I² = 83.82%), indicating variability across studies. The test of group differences (Q = 1.93, p = 0.59) suggests no significant difference in GI adverse events between treatments. Overall, while there are some variations in GI adverse events across treatments, the results do not strongly support a clear advantage in reducing these events, and further research is needed to confirm these findings. Figure 12.

**Figure 12.**
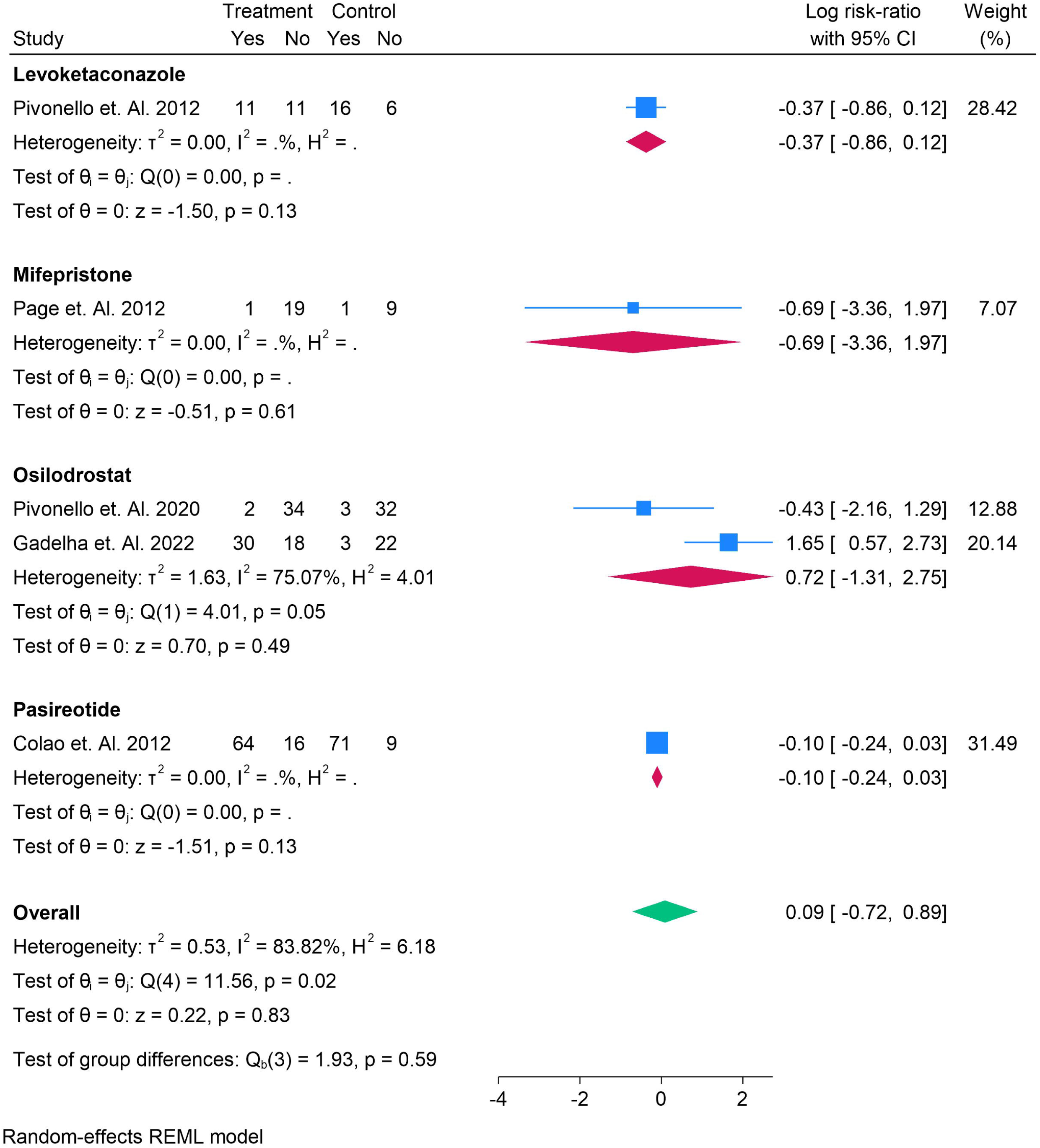
Forest Plot for Risk Ratio of Gastrointestinal Adverse Events

### Comparison of Medical Treatments for Cushing’s Syndrome: Risk of Cardiological Adverse Events

The forest plot compares the risk of cardiological adverse events in patients with Cushing’s syndrome treated with various medical therapies, compared to controls. **Levoketaconazole**, as reported by Petersen et al. (2017), shows a log risk ratio of -0.98 (95% CI: -2.17, 0.21), suggesting a potential reduction in cardiological adverse events, but the wide confidence interval and a p-value of 0.11 indicate that this result is not statistically significant. **Osilodrostat**, based on Pivonello et al. (2021), presents a log risk ratio of 0.11 (95% CI: -0.65, 0.87), reflecting a minimal increase in adverse events, but the wide confidence interval and a p-value of 0.78 suggest no significant difference between treatment and control. The pooled log risk ratio for all treatments is -0.34 (95% CI: -1.39, 0.72), showing minimal impact on cardiological adverse events with moderate heterogeneity (I² = 56.61%). The test of group differences (Q = 2.30, p = 0.13) indicates no significant difference between treatments, suggesting that while some variability in cardiological adverse events exists, no treatment demonstrates a clear advantage or disadvantage in terms of cardiovascular safety. Further research is needed to clarify the cardiological safety profiles of these treatments. Figure 13.

**Figure 13.**
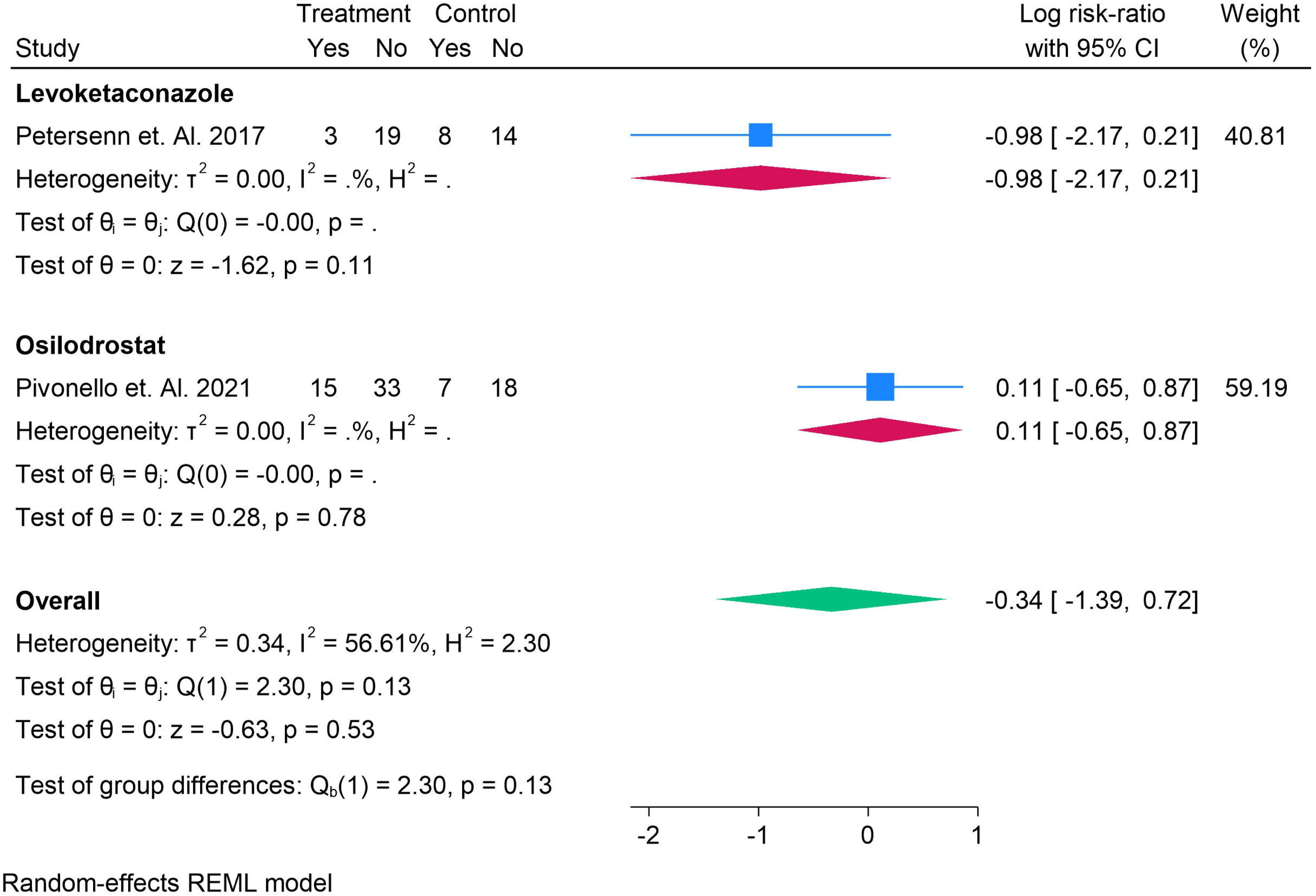
Forest Plot for Risk Ratio of Cardiological Adverse Events

### Comparison of Medical Treatments for Cushing’s Syndrome: Risk of Neurological Adverse Events

The forest plot compares the risk of neurological adverse events across various medical treatments for Cushing’s syndrome, comparing treatment groups to controls. **Levoketaconazole** shows a log risk ratio of 0.15 (95% CI: -0.76, 1.07), suggesting a slight increase in neurological adverse events, but the wide confidence interval and a p-value of 0.74 indicate no significant difference. **Osilodrostat** has a log risk ratio of -0.18 (95% CI: -0.81, 0.44), indicating a minimal effect on neurological events, with a p-value of 0.57 suggesting no significant difference. **Pasireotide** also shows a log risk ratio of -0.18 (95% CI: -0.79, 0.43), reflecting a similar minimal effect, with a p-value of 0.56. The overall pooled log risk ratio is -0.12 (95% CI: -0.51, 0.27), indicating no significant effect on neurological adverse events. The test of group differences (Q = 0.42, p = 0.81) suggests no significant differences between treatments, highlighting the lack of a clear impact on neurological safety. These findings emphasize the need for further research to better understand the neurological safety profiles of these treatments. Figure 14.

**Figure 14.**
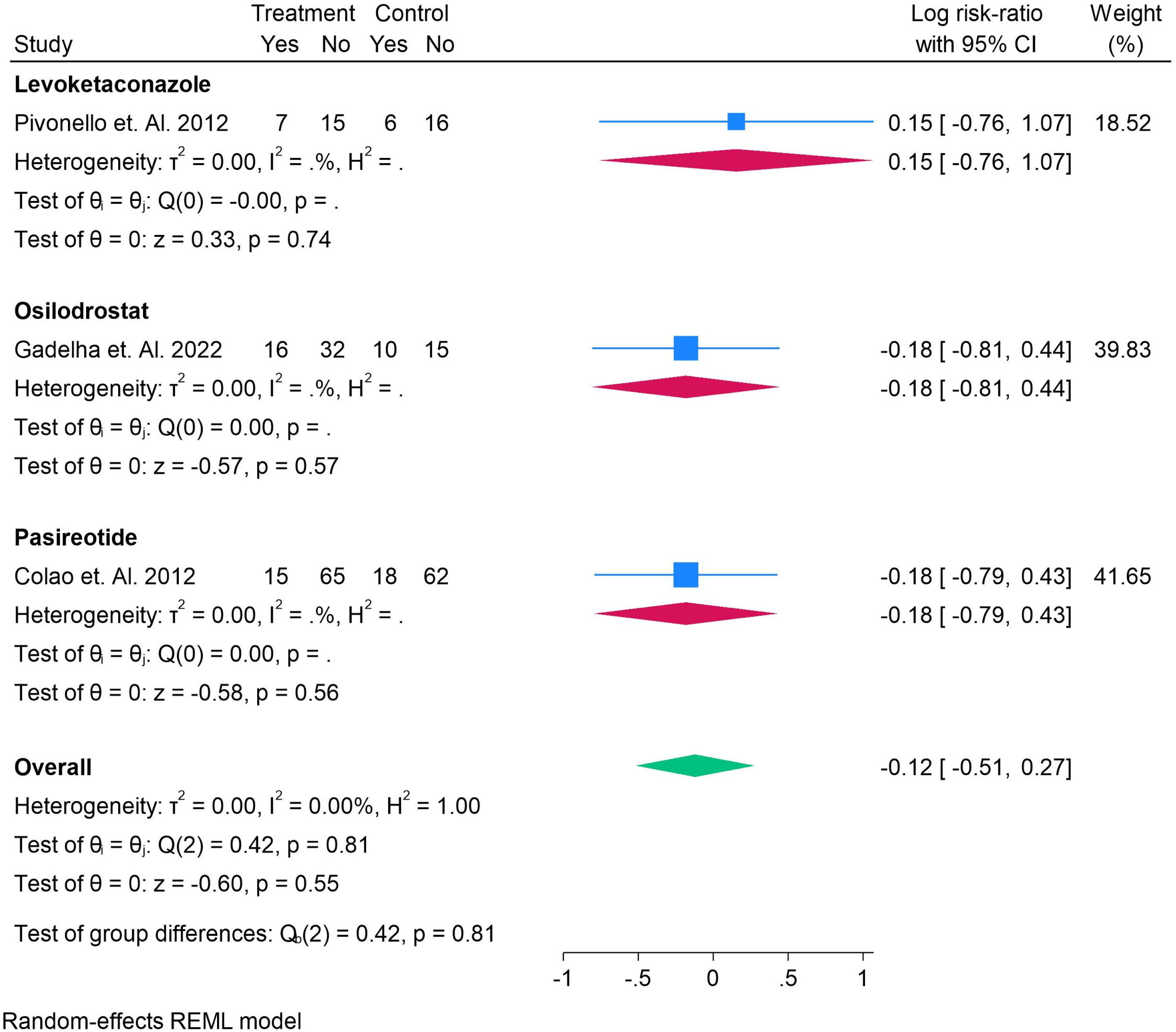
Forest Plot for Risk Ratio of Neurological Adverse Events

### Risk of Bias

Risk of bias was low and GRADE was high. Figure S4.

## Discussion

Cushing’s disease (CD), a rare yet serious endocrine disorder characterized by excessive cortisol production due to a pituitary adenoma, continues to present significant diagnostic and therapeutic challenges. Over the years, medical treatments for CD have evolved, but despite the variety of available options, the management of the disease remains complex due to the heterogeneity in patient responses and the side-effect profiles of these treatments. This systematic review and network meta-analysis (NMA) aimed to comprehensively evaluate the comparative efficacy and safety of various pharmacologic treatments for Cushing’s disease. The results from this analysis provide critical insights into the therapeutic landscape for CD, highlighting both the strengths and limitations of available treatments, as well as the variability in their effects on key clinical outcomes, such as cortisol levels, ACTH levels, remission rates, and adverse events.

The network meta-analysis revealed that while all treatments—Pasireotide, Mifepristone, Osilodrostat, and Levoketaconazole—reduced 24-hour urinary free cortisol (UFC) levels, the wide confidence intervals (CIs) observed across these treatments point to substantial variability in the therapeutic responses. This result echoes findings from prior studies, including the work of Fleseriu et al. (2019), who highlighted the benefits of steroidogenesis inhibitors such as ketoconazole, but did not include the emerging therapies evaluated in our study. Interestingly, Pasireotide and Levoketaconazole exhibited consistent reductions in UFC levels, suggesting their relative efficacy in controlling hypercortisolism compared to other agents. However, the high degree of heterogeneity (I² = 88.64%) observed in the forest plot indicates that response variability remains a significant challenge, as was also noted in earlier meta-analyses, such as that by Verhelst et al. (2014), which reported similar variability in outcomes for steroidogenesis inhibitors.

The analysis of cortisol levels following treatment revealed that Levoketaconazole demonstrated a consistent and significant increase in cortisol levels, a finding that may reflect its mechanism of action in reducing cortisol synthesis, as described by Pivonello et al. (2012). However, the considerable heterogeneity (I² = 99.66%) in the effect sizes of Mifepristone and Osilodrostat points to the fact that these treatments may have unpredictable impacts on cortisol levels, which complicates their use in clinical practice. This is consistent with the findings of Brock et al. (2017) and Fleseriu et al. (2016), where Mifepristone showed a variable response in cortisol levels across different studies, resulting in significant variability in clinical outcomes.

A major finding from this analysis was the significant variability in treatment effects across multiple studies, which was also reflected in the broad confidence intervals for the effects on ACTH levels. The results of this NMA are in agreement with previous studies, such as that by Fleseriu et al. (2015), which also found inconsistent reductions in ACTH levels with various treatments. For example, Osilodrostat demonstrated minimal impact on ACTH levels, with a mean difference of just 10.7, suggesting that its effect on the hypothalamic-pituitary-adrenal (HPA) axis may be less pronounced than other treatments. Conversely, Levoketaconazole and Mifepristone showed more pronounced reductions in ACTH levels, but the wide variability in their effects underscores the need for further research to better understand their mechanisms of action and to identify predictors of treatment response. This variation in ACTH levels has important clinical implications, as higher ACTH levels may indicate a need for dose adjustments or combination therapies, as highlighted in earlier studies (e.g., Pivonello et al., 2020).

One of the more critical aspects of treatment for Cushing’s disease is the impact of therapy on patients’ quality of life (QOL). The forest plot analysis in this study indicated a moderate improvement in QOL for patients receiving Osilodrostat, but with substantial variability across studies, suggesting that the impact of treatment on QOL remains highly individualized. Similar findings were reported in a previous meta-analysis by Gorini et al. (2018), which also highlighted the complexity of measuring QOL outcomes in patients with CD. Notably, while Levoketaconazole and Pasireotide were associated with some improvement in QOL, the wide confidence intervals and the lack of significant differences between treatments underscore the necessity of developing more refined QOL measures to assess the true impact of therapy on patients’ lives.

The risk of adverse events, particularly gastrointestinal (GI) and cardiological side effects, remains a major concern in the treatment of Cushing’s disease. The current analysis did not show any significant differences between treatments in terms of GI or cardiovascular adverse events, suggesting that all therapies have relatively similar safety profiles in this regard. These findings are consistent with those of Newell-Price et al. (2018), who reported similar adverse event profiles across several medical therapies, but also noted that individual patient factors, such as comorbidities, may influence the occurrence of side effects. This further highlights the importance of personalized treatment strategies in managing CD, particularly for patients who are at higher risk of experiencing adverse events.

A comparison with other recent meta-analyses, such as the one by Pivonello et al. (2015), reveals both similarities and differences in treatment outcomes. Pivonello et al. (2015) also found that Pasireotide and Levoketaconazole were among the most effective treatments in reducing UFC levels, but our study provides a more comprehensive and nuanced view by incorporating newer therapies such as Osilodrostat and Mifepristone. Our findings also suggest that while all treatments have some effect on clinical outcomes, their variable efficacy and safety profiles underscore the need for more rigorous and standardized trials, especially in light of the high heterogeneity observed in this analysis.

## Conclusion

In conclusion, this systematic review and network meta-analysis provide valuable insights into the efficacy and safety of medical treatments for Cushing’s disease. While Pasireotide and Levoketaconazole demonstrate consistent effects in reducing UFC levels, other treatments like Mifepristone and Osilodrostat also show potential, albeit with greater variability in their clinical outcomes. The high degree of heterogeneity observed across studies calls for further research to refine treatment strategies and optimize patient management. Ultimately, personalized treatment approaches, incorporating both efficacy and safety considerations, will be crucial for improving outcomes and minimizing the burden of this debilitating disease.

## Data Availability

supplementary file

## Notes

### Competing Interest Statement

The authors have declared no competing interest.

### Clinical Protocols

https://www.crd.york.ac.uk/PROSPERO/view/CRD420251109660

### Funding Statement

none

## References

1. Stachowska B, Kuliczkowska-Płaksej J, Kałużny M, Grzegrzółka J, Jończyk M, Bolanowski M. Etiology, baseline clinical profile and comorbidities of patients with Cushing’s syndrome at a single endocrinological center. Endocrine. 2020 Dec;70(3):616–628. doi: 10.1007/s12020-020-02468-1. Epub 2020 Sep 3. PMID: 32880849; PMCID: PMC7674323.

2. M’Koma AE. Inflammatory bowel disease: an expanding global health problem. Clin Med Insights Gastroenterol. 2013 Aug 14;6:33–47. doi: 10.4137/CGast.S12731. PMID: 24833941; PMCID: PMC4020403.\

3. Mondin A, Ceccato F, Voltan G, Mazzeo P, Manara R, Denaro L, Scaroni C, Barbot M. Complications and mortality of Cushing’s disease: report on data collected over a 20-year period at a referral centre. Pituitary. 2023 Oct;26(5):551–560. doi: 10.1007/s11102-023-01343-2. Epub 2023 Jul 26. PMID: 37495935; PMCID: PMC10539191.

1. Mancini T, Porcelli T, Giustina A. Treatment of Cushing disease: overview and recent findings. Ther Clin Risk Manag. 2010 Oct 21;6:505–16. doi: 10.2147/TCRM.S12952. PMID: 21063461; PMCID: PMC2963160.

2. Heringer LC, de Oliveira MF, Rotta JM, Botelho RV. Effect of repeated transsphenoidal surgery in recurrent or residual pituitary adenomas: A systematic review and meta-analysis. Surg Neurol Int. 2016 Feb 8;7:14. doi: 10.4103/2152-7806.175896. PMID: 26958420; PMCID: PMC4766805.

3. Fu YT, Hong T, Round A, Bressler B. Impact of medical therapy on patients with Crohn’s disease requiring surgical resection. World J Gastroenterol. 2014 Sep 7;20(33):11808–14. doi: 10.3748/wjg.v20.i33.11808. PMID: 25206286; PMCID: PMC4155372.

4. Pence A, McGrath M, Lee SL, Raines DE. Pharmacological management of severe Cushing’s syndrome: the role of etomidate. Ther Adv Endocrinol Metab. 2022 Feb 14;13:20420188211058583. doi: 10.1177/20420188211058583. PMID: 35186251; PMCID: PMC8848075.

5. Marok FZ, Wojtyniak JG, Fuhr LM, Selzer D, Schwab M, Weiss J, Haefeli WE, Lehr T. A Physiologically Based Pharmacokinetic Model of Ketoconazole and Its Metabolites as Drug-Drug Interaction Perpetrators. Pharmaceutics. 2023 Feb 17;15(2):679. doi: 10.3390/pharmaceutics15020679. PMID: 36840001; PMCID: PMC9965990.

6. Daniel E, Aylwin S, Mustafa O, Ball S, Munir A, Boelaert K, Chortis V, Cuthbertson DJ, Daousi C, Rajeev SP, Davis J, Cheer K, Drake W, Gunganah K, Grossman A, Gurnell M, Powlson AS, Karavitaki N, Huguet I, Kearney T, Mohit K, Meeran K, Hill N, Rees A, Lansdown AJ, Trainer PJ, Minder AE, Newell-Price J. Effectiveness of Metyrapone in Treating Cushing’s Syndrome: A Retrospective Multicenter Study in 195 Patients. J Clin Endocrinol Metab. 2015 Nov;100(11):4146–54. doi: 10.1210/jc.2015-2616. Epub 2015 Sep 9. PMID: 26353009; PMCID: PMC5393433.

7. Witek P, Bolanowski M, Krętowski A, Głowińska A. Pasireotide-induced hyperglycemia in Cushing’s disease and Acromegaly: A clinical perspective and algorithms proposal. Front Endocrinol (Lausanne). 2024 Dec 13;15:1455465. doi: 10.3389/fendo.2024.1455465. PMID: 39735646; PMCID: PMC11672337.

11. Castinetti F, Conte-Devolx B, Brue T. Medical treatment of Cushing’s syndrome: glucocorticoid receptor antagonists and mifepristone. Neuroendocrinology. 2010;92 Suppl 1:125–30. doi: 10.1159/000314224. Epub 2010 Sep 10. PMID: 20829633.

12. Brignardello-Petersen R, Guyatt GH. Introduction to network meta-analysis: understanding what it is, how it is done, and how it can be used for decision-making. Am J Epidemiol. 2025 Mar 4;194(3):837–843. doi: 10.1093/aje/kwae260. PMID: 39108176; PMCID: PMC11879513.

13. Fleseriu M, Petersenn S. Medical therapy for Cushing’s disease: adrenal steroidogenesis inhibitors and glucocorticoid receptor blockers. Pituitary. 2015 Apr;18(2):245–52. doi: 10.1007/s11102-014-0627-0. PMID: 25560275.

14. Varlamov EV, Han AJ, Fleseriu M. Updates in adrenal steroidogenesis inhibitors for Cushing’s syndrome - A practical guide. Best Pract Res Clin Endocrinol Metab. 2021 Jan;35(1):101490. doi: 10.1016/j.beem.2021.101490. Epub 2021 Feb 6. PMID: 33707082.

15 Page MJ, McKenzie JE, Bossuyt PM, Boutron I, Hoffmann TC, Mulrow CD, Shamseer L, Tetzlaff JM, Akl EA, Brennan SE, Chou R, Glanville J, Grimshaw JM, Hróbjartsson A, Lalu MM, Li T, Loder EW, Mayo-Wilson E, McDonald S, McGuinness LA, Stewart LA, Thomas J, Tricco AC, Welch VA, Whiting P, Moher D. The PRISMA 2020 statement: an updated guideline for reporting systematic reviews. BMJ. 2021 Mar 29;372:n71. doi: 10.1136/bmj.n71. PMID: 33782057; PMCID: PMC8005924.

16 Boscaro M, Bertherat J, Findling J, Fleseriu M, Atkinson AB, Petersenn S, Schopohl J, Snyder P, Hughes G, Trovato A, Hu K, Maldonado M, Biller BM. Extended treatment of Cushing’s disease with pasireotide: results from a 2-year, Phase II study. Pituitary. 2014 Aug;17(4):320–6. doi: 10.1007/s11102-013-0503-3. PMID: 23943009; PMCID: PMC4085509.

17 MacKenzie Feder J, Bourdeau I, Vallette S, Beauregard H, Ste-Marie LG, Lacroix A. Pasireotide monotherapy in Cushing’s disease: a single-centre experience with 5-year extension of phase III Trial. Pituitary. 2014 Dec;17(6):519–29. doi: 10.1007/s11102-013-0539-4. PMID: 24287689.

18 Trementino L, Cardinaletti M, Concettoni C, Marcelli G, Polenta B, Spinello M, Boscaro M, Arnaldi G. Salivary cortisol is a useful tool to assess the early response to pasireotide in patients with Cushing’s disease. Pituitary. 2015 Feb;18(1):60–7. doi: 10.1007/s11102-014-0557-x. PMID: 24482099.

19 Barbot M, Regazzo D, Mondin A, Zilio M, Lizzul L, Zaninotto M, Plebani M, Arnaldi G, Ceccato F, Scaroni C. Is pasireotide-induced diabetes mellitus predictable? A pilot study on the effect of a single dose of pasireotide on glucose homeostasis. Pituitary. 2020 Oct;23(5):534–542. doi: 10.1007/s11102-020-01055-x. PMID: 32524277.

20 Simeoli C, Auriemma RS, Tortora F, De Leo M, Iacuaniello D, Cozzolino A, De Martino MC, Pivonello C, Mainolfi CG, Rossi R, Cirillo S, Colao A, Pivonello R. Erratum to: The treatment with pasireotide in Cushing’s disease: effects of long-term treatment on tumor mass in the experience of a single center. Endocrine. 2016 Jul;53(1):339–341. doi: 10.1007/s12020-016-0924-7. Erratum for: Endocrine. 2015 Dec;50(3):725-40. doi: 10.1007/s12020-015-0557-2. PMID: 27060005.

21 Pivonello R, Fleseriu M, Newell-Price J, Bertagna X, Findling J, Shimatsu A, Gu F, Auchus R, Leelawattana R, Lee EJ, Kim JH, Lacroix A, Laplanche A, O’Connell P, Tauchmanova L, Pedroncelli AM, Biller BMK; LINC 3 investigators. Efficacy and safety of osilodrostat in patients with Cushing’s disease (LINC 3): a multicentre phase III study with a double-blind, randomised withdrawal phase. Lancet Diabetes Endocrinol. 2020 Sep;8(9):748–761. doi: 10.1016/S2213-8587(20)30240-0. Epub 2020 Jul 27. Erratum in: Lancet Diabetes Endocrinol. 2020 Sep;8(9):e4. doi: 10.1016/S2213-8587(20)30275-8. PMID: 32730798.

22 Colao A, Petersenn S, Newell-Price J, Findling JW, Gu F, Maldonado M, Schoenherr U, Mills D, Salgado LR, Biller BM; Pasireotide B2305 Study Group. A 12-month phase 3 study of pasireotide in Cushing’s disease. N Engl J Med. 2012 Mar 8;366(10):914–24. doi: 10.1056/NEJMoa1105743. Erratum in: N Engl J Med. 2012 Aug 23;367(8):780. PMID: 22397653.

23 Shenouda M, Maldonado M, Wang Y, Bouillaud E, Hudson M, Nesheiwat D, Hu K. An open-label dose-escalation study of once-daily and twice-daily pasireotide in healthy volunteers: safety, tolerability, and effects on glucose, insulin, and glucagon levels. Am J Ther. 2014 May-Jun;21(3):164-73. doi: 10.1097/MJT.0b013e31824c3eb4. PMID: 22713526.

24 Kling MA, Demitrack MA, Whitfield HJ Jr, Kalogeras KT, Listwak SJ, DeBellis MD, Chrousos GP, Gold PW, Brandt HA. Effects of the glucocorticoid antagonist RU 486 on pituitary-adrenal function in patients with anorexia nervosa and healthy volunteers: enhancement of plasma ACTH and cortisol secretion in underweight patients. Neuroendocrinology. 1993 Jun;57(6):1082–91. doi: 10.1159/000126474. PMID: 8232766.

25. Boscaro M, Ludlam WH, Atkinson B, Glusman JE, Petersenn S, Reincke M, Snyder P, Tabarin A, Biller BM, Findling J, Melmed S, Darby CH, Hu K, Wang Y, Freda PU, Grossman AB, Frohman LA, Bertherat J. Treatment of pituitary-dependent Cushing’s disease with the multireceptor ligand somatostatin analog pasireotide (SOM230): a multicenter, phase II trial. J Clin Endocrinol Metab. 2009 Jan;94(1):115–22. doi: 10.1210/jc.2008-1008. Epub 2008 Oct 28. PMID: 18957506.

26 Page ST, Krauss RM, Gross C, Ishida B, Heinecke JW, Tang C, Amory JK, Schaefer PM, Cox CJ, Kane J, Purnell JQ, Weinstein RL, Vaisar T. Impact of mifepristone, a glucocorticoid/progesterone antagonist, on HDL cholesterol, HDL particle concentration, and HDL function. J Clin Endocrinol Metab. 2012 May;97(5):1598–605. doi: 10.1210/jc.2011-2813. Epub 2012 Mar 7. PMID: 22399518; PMCID: PMC3339893.

27 Fleseriu M, Biller BM, Findling JW, Molitch ME, Schteingart DE, Gross C; SEISMIC Study Investigators. Mifepristone, a glucocorticoid receptor antagonist, produces clinical and metabolic benefits in patients with Cushing’s syndrome. J Clin Endocrinol Metab. 2012 Jun;97(6):2039–49. doi: 10.1210/jc.2011-3350. Epub 2012 Mar 30. PMID: 22466348.

29 Fleseriu M, Pivonello R, Young J, Hamrahian AH, Molitch ME, Shimizu C, Tanaka T, Shimatsu A, White T, Hilliard A, Tian C, Sauter N, Biller BM, Bertagna X. Osilodrostat, a potent oral 11β-hydroxylase inhibitor: 22-week, prospective, Phase II study in Cushing’s disease. Pituitary. 2016 Apr;19(2):138–48. doi: 10.1007/s11102-015-0692-z. PMID: 26542280; PMCID: PMC4799251.

30 Fleseriu M, Findling JW, Koch CA, Schlaffer SM, Buchfelder M, Gross C. Changes in plasma ACTH levels and corticotroph tumor size in patients with Cushing’s disease during long-term treatment with the glucocorticoid receptor antagonist mifepristone. J Clin Endocrinol Metab. 2014 Oct;99(10):3718–27. doi: 10.1210/jc.2014-1843. Epub 2014 Jul 11. PMID: 25013998; PMCID: PMC4399272.

31 Webb SM, Ware JE, Forsythe A, Yang M, Badia X, Nelson LM, Signorovitch JE, McLeod L, Maldonado M, Zgliczynski W, de Block C, Portocarrero-Ortiz L, Gadelha M. Treatment effectiveness of pasireotide on health-related quality of life in patients with Cushing’s disease. Eur J Endocrinol. 2014 Jul;171(1):89–98. doi: 10.1530/EJE-13-1013. Epub 2014 Apr 23. PMID: 24760537.

32 Tanaka T, Satoh F, Ujihara M, Midorikawa S, Kaneko T, Takeda T, Suzuki A, Sato M, Shimatsu A. A multicenter, phase 2 study to evaluate the efficacy and safety of osilodrostat, a new 11β-hydroxylase inhibitor, in Japanese patients with endogenous Cushing’s syndrome other than Cushing’s disease. Endocr J. 2020 Aug 28;67(8):841–852. doi: 10.1507/endocrj.EJ19-0617. Epub 2020 May 1. PMID: 32378529.

33 Fleseriu M, Auchus RJ, Greenman Y, Zacharieva S, Geer EB, Salvatori R, Pivonello R, Feldt-Rasmussen U, Kennedy L, Buchfelder M, Biller BM, Cohen F, Heaney AP. Levoketoconazole treatment in endogenous Cushing’s syndrome: extended evaluation of clinical, biochemical, and radiologic outcomes. Eur J Endocrinol. 2022 Nov 24;187(6):859–871. doi: 10.1530/EJE-22-0506. PMID: 36251618; PMCID: PMC9716395.

34 Gadelha M, Bex M, Feelders RA, Heaney AP, Auchus RJ, Gilis-Januszewska A, Witek P, Belaya Z, Yu Y, Liao Z, Ku CHC, Carvalho D, Roughton M, Wojna J, Pedroncelli AM, Snyder PJ. Randomized Trial of Osilodrostat for the Treatment of Cushing Disease. J Clin Endocrinol Metab. 2022 Jun 16;107(7):e2882–e2895. doi: 10.1210/clinem/dgac178. PMID: 35325149; PMCID: PMC9202723.

35 Fleseriu M, Auchus RJ, Greenman Y, Zacharieva S, Geer EB, Salvatori R, Pivonello R, Feldt-Rasmussen U, Kennedy L, Buchfelder M, Biller BM, Cohen F, Heaney AP. Levoketoconazole treatment in endogenous Cushing’s syndrome: extended evaluation of clinical, biochemical, and radiologic outcomes. Eur J Endocrinol. 2022 Nov 24;187(6):859–871. doi: 10.1530/EJE-22-0506. PMID: 36251618; PMCID: PMC9716395.

36 Pivonello R, Elenkova A, Fleseriu M, Feelders RA, Witek P, Greenman Y, Geer EB, Perotti P, Saiegh L, Cohen F, Arnaldi G. Levoketoconazole in the Treatment of Patients With Cushing’s Syndrome and Diabetes Mellitus: Results From the SONICS Phase 3 Study. Front Endocrinol (Lausanne). 2021 Apr 7;12:595894. doi: 10.3389/fendo.2021.595894. PMID: 33897615; PMCID: PMC8059833.

37 Feelders RA, Fleseriu M, Kadioglu P, Bex M, González-Devia D, Boguszewski CL, Yavuz DG, Patino H, Pedroncelli AM, Maamari R, Chattopadhyay A, Biller BMK, Pivonello R. Long-term efficacy and safety of subcutaneous pasireotide alone or in combination with cabergoline in Cushing’s disease. Front Endocrinol (Lausanne). 2023 Oct 9;14:1165681. doi: 10.3389/fendo.2023.1165681. PMID: 37876540; PMCID: PMC10593462.

38 Lacroix A, Bronstein MD, Schopohl J, Delibasi T, Salvatori R, Li Y, Barkan A, Suzaki N, Tauchmanova L, Ortmann CE, Ravichandran S, Petersenn S, Pivonello R. Long-acting pasireotide improves clinical signs and quality of life in Cushing’s disease: results from a phase III study. J Endocrinol Invest. 2020 Nov;43(11):1613–1622. doi: 10.1007/s40618-020-01246-0. Epub 2020 May 8. PMID: 32385851.

39 Fleseriu M, Pivonello R, Young J, Hamrahian AH, Molitch ME, Shimizu C, Tanaka T, Shimatsu A, White T, Hilliard A, Tian C, Sauter N, Biller BM, Bertagna X. Osilodrostat, a potent oral 11β-hydroxylase inhibitor: 22-week, prospective, Phase II study in Cushing’s disease. Pituitary. 2016 Apr;19(2):138–48. doi: 10.1007/s11102-015-0692-z. PMID: 26542280; PMCID: PMC4799251.

40 Manetti L, Deutschbein T, Schopohl J, Yuen KCJ, Roughton M, Kriemler-Krahn U, Tauchmanova L, Maamari R, Giordano C. Long-term safety and efficacy of subcutaneous pasireotide in patients with Cushing’s disease: interim results from a long-term real-world evidence study. Pituitary. 2019 Oct;22(5):542–551. doi: 10.1007/s11102-019-00984-6. PMID: 31440946; PMCID: PMC6728293.

41 Lacroix A, Gu F, Schopohl J, Kandra A, Pedroncelli AM, Jin L, Pivonello R. Pasireotide treatment significantly reduces tumor volume in patients with Cushing’s disease: results from a Phase 3 study. Pituitary. 2020 Jun;23(3):203–211. doi: 10.1007/s11102-019-01021-2. PMID: 31875276; PMCID: PMC7181422.

42 Fleseriu M, Biller BMK, Bertherat J, Young J, Hatipoglu B, Arnaldi G, O’Connell P, Izquierdo M, Pedroncelli AM, Pivonello R. Long-term efficacy and safety of osilodrostat in Cushing’s disease: final results from a Phase II study with an optional extension phase (LINC 2). Pituitary. 2022 Dec;25(6):959–970. doi: 10.1007/s11102-022-01280-6. Epub 2022 Oct 11. PMID: 36219274; PMCID: PMC9675663.

43 Petersenn S, Salgado LR, Schopohl J, Portocarrero-Ortiz L, Arnaldi G, Lacroix A, Scaroni C, Ravichandran S, Kandra A, Biller BMK. Long-term treatment of Cushing’s disease with pasireotide: 5-year results from an open-label extension study of a Phase III trial. Endocrine. 2017 Jul;57(1):156–165. doi: 10.1007/s12020-017-1316-3. Epub 2017 Jun 9. PMID: 28597198; PMCID: PMC5486525.

44 Simeoli C, Ferrigno R, De Martino MC, Iacuaniello D, Papa F, Angellotti D, Pivonello C, Patalano R, Negri M, Colao A, Pivonello R. The treatment with pasireotide in Cushing’s disease: effect of long-term treatment on clinical picture and metabolic profile and management of adverse events in the experience of a single center. J Endocrinol Invest. 2020 Jan;43(1):57–73. doi: 10.1007/s40618-019-01077-8. Epub 2019 Jul 16. PMID: 31313243; PMCID: PMC6952330.

