## supplementary file for "Comparative Efficacy and Safety of Medical Treatments for Cushing’s Disease: A Systematic Review and Network Meta-Analysis"

Supplementary Files

Search String

("Cushing's disease" OR "Cushing syndrome") AND ("medical treatments" OR "treatment" OR "therapy" OR "medications") AND ("efficacy" OR "effectiveness" OR "clinical outcomes" OR "treatment response") AND ("safety" OR "adverse events" OR "side effects")

Table 1. Demographics and Summary of findings

| **Ref Num** | **Author Year** | **Country** | **Total Sample** | **Male** | **Female** | **Age** | **Follow Up Time** | **Treatment Name** | **Sample in treatment** | **Control name** | **Sample in Treatment** | **GRADE** | **Main Finding** |
| --- | --- | --- | --- | --- | --- | --- | --- | --- | --- | --- | --- | --- | --- |
| 16 | Boscaro et. Al. 2013 | Multicenter | 19 | 2 | 17 | 43 | 9.7 | Pasireotide | 19 | Single Arm Study | N/A | High | The study suggests that **Pasireotide is effective for long-term treatment of Cushing’s disease, offering sustained reductions in UFC levels and clinical improvements** |
| 17 | Feder et. Al.2013 | Canada | 4 | 0 | 4 | 52 | 50.4 | Pasireotide | 4 | Single Arm Study | N/A | High | Pasireotide therapy can provide sustained normalization of UFC and clinical remission in Cushing’s disease over long-term follow-up |
| 18 | Trementino et. Al. 2014 | Italy | 7 | 2 | 5 | 35.3 | 7 | Pasireotide | 7 | Single Arm Study | N/A | High | Pasireotide rapidly reduced and normalized both UFC and salivary cortisol levels in patients with Cushing’s disease |
| 19 | Barbot et. Al. 2020 | Italy | 15 | 2 | 13 | 43 | 6 | Pasireotide | 15 | Single Arm Study | N/A | High | Pasireotide (PAS) rapidly impacts glucose metabolism and may induce diabetes in patients with Cushing's disease, especially those with higher glucagon and HbA1c levels |
| 20 | Pivonello et. Al. 2014 | Italy | 8 | 1 | 7 | 38.9 | 12 | Pasireotide | 8 | Single Arm Study | N/A | High | The study found that Pasireotide treatment significantly reduced UFC levels and tumor volume in patients with Cushing's disease |
| 21 | Pivonello et. Al. 2020 | Multicenter | 137 | 31 | 106 | 40 | 74.7 | Osilodrostat | 36 | Placebo | 35 | High | Osilodrostat is an **effective treatment for Cushing's disease**, with a high rate of complete response compared to placebo |
| 22 | Colao et. Al. 2012 | Multicenter | 162 | 32 | 126 | 40 | 10.8 | Pasireotide | 80 | Placebo | 80 | High | Pasireotide significantly reduced cortisol levels in Cushing's disease patients, showing its potential for use as a targeted therapy for corticotropin-secreting pituitary adenomas. |
| 23 | Shenouda et. Al. 2014 | USA | 66 | 66 | 0 | 26 | 0.25 | Pasireotide | 66 | Single Arm Study | N/A | High | Pasireotide showed good overall tolerability up to doses of 1500 mg once daily and 750 mg twice daily for 8 days, with fasting and postprandial hyperglycemia occurring in all participants |
| 24 | Kling et. al. 1993 | USA | 15 | 0 | 15 | 23.5 | 1.5 | Mifepristone | 8 | Placebo | 8 | High | Hypercortisolism in underweight anorexia nervosa patients reflects hypersecretion of hypothalamic CRH, not glucocorticoid resistance, with persistent adrenal hyperresponsiveness post-weight recovery. |
| 25 | Boscaro et. Al. 2009 | USA | 39 | 29 | 10 | 41.5 | 15 | Pasireotide | 39 | Single Arm Study | N/A | High | Pasireotide reduced UFC levels in 76% of Cushing’s disease patients, with 17% achieving normalization, showing promise as a pituitary-targeted therapy. |
| 26 | Page et. Al. 2012 | USA | 30 | 0 | 30 | 59 | 1.5 | Mifepristone | 20 | Placebo | 10 | High | Mifepristone reduced HDL-C and HDL particle concentration but improved HDL function on a per-particle basis. |
| 27 | Fleseriu et. Al. 2012 | USA | 50 | 15 | 35 | 45.5 | 6 | Mifepristone | 50 | Single Arm Study | N/A | High | Mifepristone significantly improved clinical and metabolic outcomes in patients with Cushing’s syndrome with an acceptable safety profile. |
| 28 | Bertagna et. Al. 2014 | Multicenter | 12 | 8 | 4 | 39 | 10 | Osilodrostat | 12 | Single Arm Study | N/A | Moderate | **LCI699 effectively normalized UFC levels in 92% of Cushing’s disease patients and was well tolerated**. |
| 29 | Fieseriu et. Al. 2014 | Multicenter | 43 | 11 | 32 | 45.3 | 11.3 | Mifepristone | 43 | Single Arm Study | N/A | High | Long-term mifepristone treatment in Cushing’s disease patients increased ACTH levels (72% had ≥2-fold rise) with dose-dependent effects, while tumor progression was rare (4/36 cases). |
| 30 | Bertagna et. Al. 186 | France | 7 | 3 | 4 | 35 | 0.25 | Mifepristone | 5 | Placebo | 2 | High | RU 486 induced a delayed and prolonged pituitary-adrenal response in Cushing's disease but not in nonpituitary-dependent Cushing's syndrome. |
| 31 | Webb et. Al. 2014 | Multicenter | 162 | 23 | 139 | 40.7 | 12 | Pasireotide | 162 | Single Arm Study | N/A | High | Pasireotide treatment improved HRQoL in Cushing’s disease patients, particularly those achieving biochemical control. |
| 32 | Tanaka et. Al. 2020 | Japan | 9 | 7 | 2 | 46 | 12 | Osilodrostat | 9 | Single Arm Study | N/A | High | Osilodrostat effectively reduced mUFC levels in Japanese patients with endogenous Cushing’s syndrome (non-Cushing’s disease) and had a manageable safety profile. |
| 33 | Fleseriu et. Al. 2019 | Multicenter | 81 | 20 | 61 | 39.7 | 23.9 | Pasireotide | 81 | Single Arm Study | N/A | High | Long-acting pasireotide provided sustained biochemical and clinical improvements in Cushing’s disease with a manageable safety profile. |
| 34 | Gadelha et. Al. 2022 | Multicenter | 73 | 61 | 12 | 39 | 70 | Osilodrostat | 48 | Placebo | 25 | High | **Osilodrostat rapidly normalized mUFC excretion in most patients with Cushing disease and maintained this effect with a favorable safety profile.** |
| 35 | Fleseriu et. Al. 2022 | Multicenter | 60 | 50 | 10 | 43.8 | 15 | Levoketaconazole | 60 | Single Arm Study | N/A | High | Levoketoconazole sustained clinical and biochemical benefits in Cushing’s syndrome over 12 months with no new safety concerns. |
| 36 | Pivonello et. Al. 2021 | Multicenter | 94 | 17 | 77 | 48.3 | 6 | Levoketaconazole | 94 | Single Arm Study | N/A | High | **Levoketoconazole normalized mUFC and improved glycemic control in Cushing’s syndrome patients, with pronounced benefits in diabetic patients.** |
| 37 | Feelders et. Al. 2023 | Multicenter | 68 | 8 | 60 | 41.4 | 35 | Pasireotide | 26 | Placebo | 42 | High | **Combination therapy with pasireotide and cabergoline effectively enhanced long-term control of hypercortisolism in Cushing’s disease patients, with a tolerable safety profile.** |
| 38 | Lacroix et. Al. 2020 | Multicenter | 150 | 118 | 32 | 38.5 | 12 | Pasireotide | 150 | Single Arm Study | N/A | High | Long-acting pasireotide improved clinical signs and HRQoL in Cushing’s disease, with benefits seen even in patients without full UFC control |
| 39 | Fleseriu et. Al. 2016 | Multicenter | 19 | 5 | 14 | 36.8 | 6.7 | Osilodrostat | 19 | Single Arm Study | N/A | High | Osilodrostat effectively normalized UFC in 78.9% of Cushing’s disease patients by week 22 and was generally well tolerated. |
| 40 | Manetti et. Al. 2019 | Multicenter | 127 | 26 | 101 | 49.9 | 14 | Pasireotide | 127 | Single Arm Study | N/A | High | Pasireotide is well tolerated and provides sustained reductions in mUFC in real-world treatment of Cushing’s disease, with manageable hyperglycemia. |
| 41 | Pivonello et. Al. 2012 | Multicenter | 84 | 20 | 64 | 44.7 | 30 | Levoketaconazole | 22 | Placebo | 22 | High | Levoketoconazole effectively normalized urinary cortisol in Cushing’s syndrome patients, with reversible effects upon withdrawal and no new safety risks identified. |
| 42 | Fleseriu et. Al. 2022b | Multicenter | 19 | 5 | 14 | 36.8 | 64.8 | Osilodrostat | 19 | Single Arm Study | N/A | High | Osilodrostat provided sustained reductions in mUFC for up to 6.7 years in Cushing’s disease patients, with no new safety signals. |
| 43 | Petersenn et. Al. 2017 | Multicenter | 162 | 36 | 126 | 40.2 | 60 | Pasireotide | 162 | Single Arm Study | N/A | High | Pasireotide provided sustained biochemical and clinical improvements in Cushing’s disease over 5 years, with a manageable safety profile. |
| 44 | Simeoli et. Al. 2020 | Italy | 14 | 2 | 12 | 39.07 | 12 | Pasireotide | 14 | Single Arm Study | N/A | High | Pasireotide effectively controlled cortisol secretion and improved clinical/metabolic parameters in Cushing’s disease patients, but required careful management of hyperglycemia and other adverse events. |

Table S2. Ranking Table for UFC in 24 hours in various medical treatments for cushing syndrome

|  | **Levoketaconazole** | **Mifepristone** | **Osilodrostat** | **Pasireotide** | **Placebo** |
| --- | --- | --- | --- | --- | --- |
| **Levoketaconazole** | **Levoketaconazole** | **-6.74 (-3836.31, 2698.89)** | **119.68 (-2325.27, 2620.46)** | **-1.66 (-2487.47, 2461.23)** | **328.64 (-1703.75, 2332.83)** |
| **Mifepristone** | **6.74 (-2698.89, 3836.31)** | **Mifepristone** | **148.62 (-2231.95, 3561.83)** | **6.25 (-2402.72, 3442.13)** | **380.89 (-1736.58, 3267.79)** |
| **Osilodrostat** | **-119.68 (-2620.46, 2325.27)** | **-148.62 (-3561.83, 2231.95)** | **Osilodrostat** | **-115.54 (-2158.53, 1861.67)** | **213.69 (-1247.29, 1600.36)** |
| **Pasireotide** | **1.66 (-2461.23, 2487.47)** | **-6.25 (-3442.13, 2402.72)** | **115.54 (-1861.67, 2158.53)** | **Pasireotide** | **331.27 (-1091.39, 1749.62)** |
| **Placebo** | **-328.64 (-2332.83, 1703.75)** | **-380.89 (-3267.79, 1736.58)** | **-213.69 (-1600.36, 1247.29)** | **-331.27 (-1749.62, 1091.39)** | **Placebo** |

Figure S1. SUCRA Ranking for 24 hours in various medical treatments for cushing syndrome

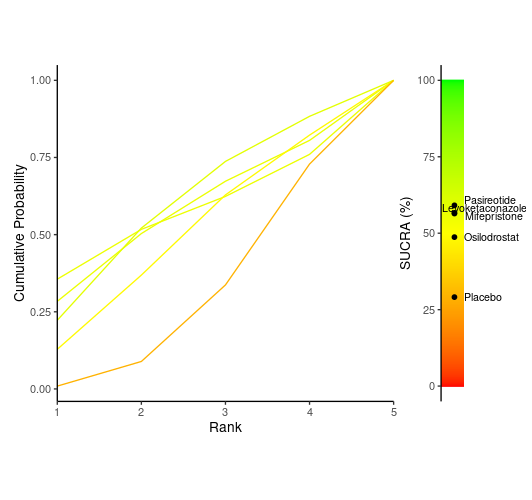

Table S3. Ranking Table for Cortisol Levels in various medical treatments for cushing syndrome

|  | **Levoketaconazole** | **Mifepristone** | **Osilodrostat** | **Placebo** |
| --- | --- | --- | --- | --- |
| **Levoketaconazole** | **Levoketaconazole** | **26.59 (-106.93, 160.55)** | **-6.52 (-141.67, 126.89)** | **-17.06 (-127.35, 91.83)** |
| **Mifepristone** | **-26.59 (-160.55, 106.93)** | **Mifepristone** | **-33.31 (-143.31, 77.68)** | **-43.6 (-121.82, 34.32)** |
| **Osilodrostat** | **6.52 (-126.89, 141.67)** | **33.31 (-77.68, 143.31)** | **Osilodrostat** | **-10.66 (-88.45, 67.35)** |
| **Placebo** | **17.06 (-91.83, 127.35)** | **43.6 (-34.32, 121.82)** | **10.66 (-67.35, 88.45)** | **Placebo** |

Table S2. SUCRA Ranking for Cortisol Levels in various medical treatments for cushing syndrome

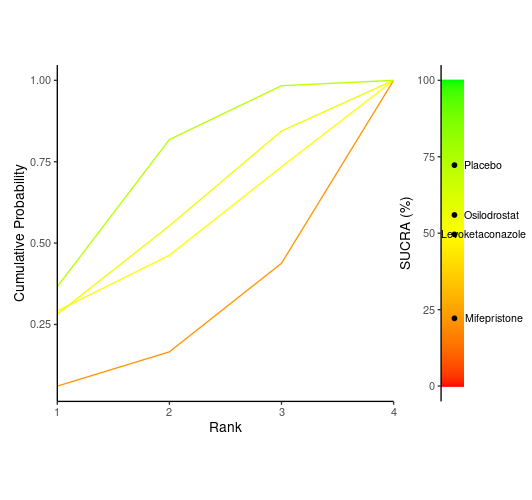

Table S4. Ranking Table for change in ACTH in various medical treatments for cushing syndrome

|  | **Levoketaconazole** | **Mifepristone** | **Osilodrostat** | **Placebo** |
| --- | --- | --- | --- | --- |
| **Levoketaconazole** | **Levoketaconazole** | **26.59 (-106.93, 160.55)** | **-6.52 (-141.67, 126.89)** | **-17.06 (-127.35, 91.83)** |
| **Mifepristone** | **-26.59 (-160.55, 106.93)** | **Mifepristone** | **-33.31 (-143.31, 77.68)** | **-43.6 (-121.82, 34.32)** |
| **Osilodrostat** | **6.52 (-126.89, 141.67)** | **33.31 (-77.68, 143.31)** | **Osilodrostat** | **-10.66 (-88.45, 67.35)** |
| **Placebo** | **17.06 (-91.83, 127.35)** | **43.6 (-34.32, 121.82)** | **10.66 (-67.35, 88.45)** | **Placebo** |

Figure S3. SUCRA Ranking for ACTH Levels in various medical treatments for cushing syndrome

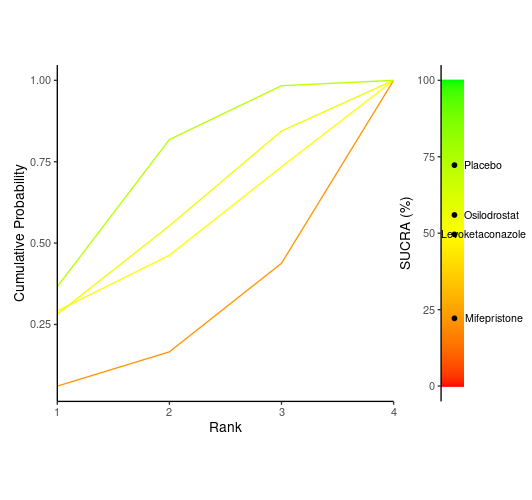

Figure S4. Risk of Bias ROBS 2.0

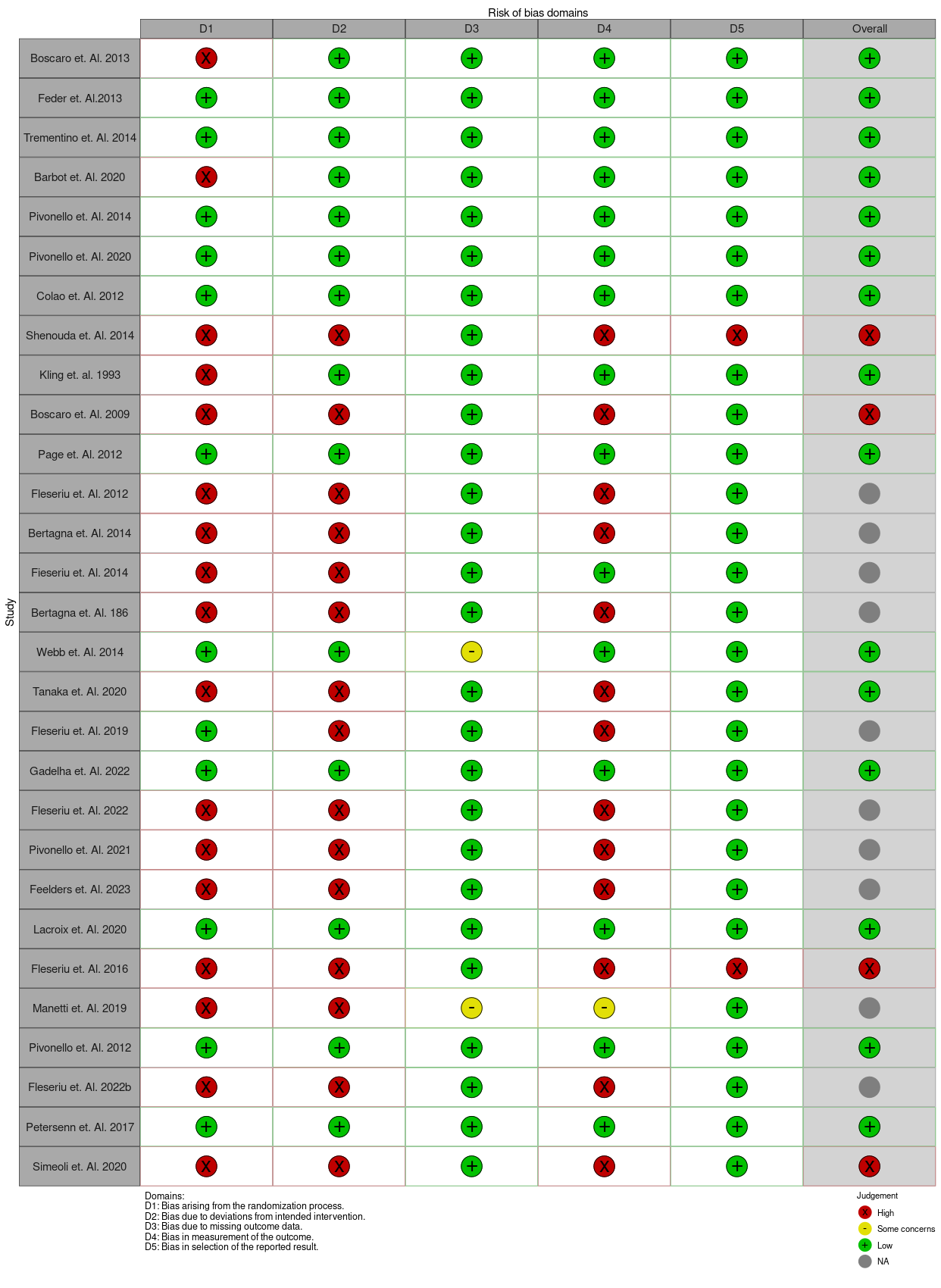
